# Disproportionate Mental Health Risks in Autistic Females: A Rapid Review with Quantitative and Narrative Syntheses

**DOI:** 10.1101/2025.09.12.25335643

**Authors:** Adeline Lacroix, Chih-Chen Tzang, Jennifer Xiaofan Yu, Benjamin Koshy Jacob, Mishel Alexandrovsky, Anna Winge-Breen, Terri Rodak, Meng-Chuan Lai

## Abstract

Mental health conditions are highly prevalent among autistic people, but an updated synthesis of sex-stratified prevalence data, contributing factors, and support strategies is lacking. To address this knowledge gap, we conducted a rapid review utilizing PRISMA-ScR guidelines. MEDLINE, CINAHL, and PsycINFO databases were searched for studies (2004–2024) including female participants with a clinical autism diagnosis, and with a focus on mental health. Of 8,420 records screened, 218 met inclusion criteria. An exploratory quantitative synthesis of population-based and registry-based studies revealed higher rates of mental health conditions in autistic females than males for anxiety, mood, eating, obsessive-compulsive, psychotic, and personality disorders. Narrative synthesis identified moderating factors, including sex-related physiology, gendered experiences, age, age at autism diagnosis, autism characteristics, and co-occurring conditions. Biological and social mechanisms likely interact as contributing factors. Severe consequences of poor mental health underscore the need for tailored approaches accounting for the specific profiles of autistic females.

## Introduction

Autism occurs in at least 0.6–1% of the global population (Salari et al., 2022; Talantseva et al., 2023; Zeidan et al., 2022) and is characterized by social-communication differences, restricted and repetitive patterns of behavior and interests, and atypical sensory responses. Autistic people (Keating et al., 2023) also experience higher rates of co-occurring mental health (MH) conditions than non-autistic people (Lai et al., 2019; Micai et al., 2023). Given the significant impact of MH on quality of life (Mason et al., 2018), enhancing MH services for autistic individuals is a top research and clinical priority (Grove et al., 2025; Wechsler et al., 2025).

MH conditions are sometimes diagnosed, or misdiagnosed (Dell’Osso & Carpita, 2023) prior to autism, potentially contributing to delays in autism diagnoses (Gu et al., 2023; Wodka et al., 2022). This pattern is especially observed in autistic females, who are often identified later than males (Gu et al., 2023; Harrop et al., 2024; Smith et al., 2024). Under-recognition contributes to the lower reported prevalence of autism in females, with a widely cited sex ratio of 1:4 female-to-male (Loomes et al., 2017; Zeidan et al., 2022). However, when using active case ascertainment methods, the estimated ratio narrows to 1:3.25 (Loomes et al., 2017; O’Nions et al., 2023). Although still imbalanced, this difference may reflect a combination of biological factors affecting the likelihood of autism expression in females (Dougherty et al., 2022; Lai et al., 2015; Leow et al., 2024), and gendered socio-cultural factors shaping the presentation and perception of autism in females (Lai et al., 2015; Lai, Amestoy, et al., 2023).

The interaction of biological sex-related and social gender-related factors may also affect the prevalence and presentation of co-occurring health conditions in autistic people. While several meta-analyses have summarized the overall prevalence of MH diagnoses (Lai et al., 2019; Mutluer et al., 2022; Sigurdardottir et al., 2025), few have systematically synthesized sex-specific profiles. Two recent narrative reviews have examined aspects of MH in autistic females, though MH was not the sole focus (Rynkiewicz et al., 2024; Tricco et al., 2018). Moreover, neither employed a systematic search strategy, and one focused on neurodivergent populations more broadly (Kelly et al., 2024). As a result, the potential factors contributing to sex and gender differences in MH outcomes in autistic people were not thoroughly examined. Overall, MH in autistic females remains relatively under-investigated and existing knowledge lacks systematic synthesis (Lai, Amestoy, et al., 2023).

Similar to our prior efforts in synthesizing the knowledge about physical health in autistic females (Kassee et al., 2020), we conducted a rapid review focusing on the MH of autistic females. Using systematic search and scoping review methodologies, we combined exploratory quantitative analysis and narrative synthesis to: (1) map sex differences in the prevalence and presentation of MH conditions in autistic people, (2) explore factors influencing these differences, (3) consider explanations for these sex differences, and (4) summarize downstream consequences and suggestions for tailored support for autistic females.

We focused on differences reported based on sex, which in most studies corresponds to a binary designation (female/male) derived from anatomical characteristics recorded at birth. Accordingly, we used the terms “female” and “male” when referring to these sex-based categories. We acknowledge the natural diversity of gender identities and the important influences of gender socialization, often imposed on individuals based on sex assigned at birth. When specifically discussing gender identity-related observations, we used the terms “assigned female/male at birth” (AFAB/AMAB). This terminology allows us to refer to sex-related categories while acknowledging that they do not necessarily reflect participants’ gender identities. We also note that the effects of socialization, and its interaction with gender identity, may be embedded in the variance observed in relation to sex-based categories.

## Method

This rapid review was guided by the Preferred Reporting Items for Systematic reviews and Meta-Analyses extension for scoping reviews (PRISMA-ScR; Tricco et al., 2018). The search strategy was developed by a health sciences research librarian (TR) in collaboration with M-CL and AL. Searches were conducted in MEDLINE, APA PsycINFO, and CINAHL on October 29, 2024. All strategies used database-specific subject headings, keywords in natural language, and advanced search operators. To exclude animal studies, a search filter from Canada’s Drug Agency was applied (Canada’s Drug Agency, 2025b, 2025a), and results were limited to publication date from January 1, 2004 to the search date. No study design or language limits were applied. The full MEDLINE search strategy is detailed in Supplementary Table1.

Peer-reviewed journal papers, including reviews, meta-analyses, commentaries, opinion pieces, editorials, letters to editors, and case studies, were included if they were written in English, involved participants with clinical diagnoses of autism, included or focused solely on autistic females, and incorporated a measurement (i.e., reported clinical diagnosis, validated dimensional measure of MH symptoms) or a discussion with a direct relevance to MH in autistic females. References to “gender” in the included studies were examined carefully to determine whether they referred to sex or gender identity, as the use of these terms had unfortunately often been conflated in past empirical research. Given the heterogeneity in how MH conditions were ascertained across studies, including the use of different classification systems (e.g., ICD-8, ICD-9, ICD-10), algorithmic recoding of diagnostic codes, and parent- or self-reports, we categorized MH conditions into broad, commonly used clinical groupings to enable meaningful synthesis. These include anxiety disorders, mood disorders (further divided into mood [affective] disorders in general, depressive disorders, and bipolar disorders), eating disorders (ED), obsessive-compulsive disorder (OCD), personality disorders, schizophrenia spectrum and other psychotic disorders, suicidality (further divided by suicidal ideation, suicide attempts and self-harm requiring hospitalization, and suicide deaths), and substance use disorders (SUD). Results for other MH conditions, for which data were limited (e.g., Oppositional Defiant Disorder, Post-Traumatic Stress Disorder, geriatric neurological conditions), are reported in the Supplementary Materials, along with findings on transdiagnostic symptoms and association patterns of co-occurring conditions. Gender dysphoria, other neurodevelopmental conditions such as attention-deficit/hyperactivity disorder (ADHD) or intellectual disabilities (ID), and sleep-wake disorders were included in the search and considered as co-occurring conditions potentially modulating or modulated by MH, rather than MH conditions per se. Papers were excluded if (1) there were no clear differentiation between self-identified autistic participants and clinically diagnosed autistic participants in the analysis; (2) analyses or discussions were not articulated in a sex-stratified manner; or (3) the paper did not refer to a MH issue.

Database search results were imported into Covidence (Veritas Health Innovation, Melbourne, Australia). Six author-reviewers (AL, C-CT, BK-J, JXY, MA, AW-B) screened titles and abstracts, with support of the senior author (M-CL), using broad inclusion criteria. A one-hour session where all reviewers screened the same abstracts was conducted to ensure calibration. Thereafter, abstracts were independently screened by single reviewers, with ambiguous cases discussed collaboratively within the team. The same procedure was applied for full-text screening. At this stage, following team discussions and given the large number of low-relevance studies, an additional screening step was implemented, applying more stringent exclusion criteria to identify papers with methodological limitations. Studies were excluded if they (4) were case-control studies with fewer than 15 autistic females and less than 12.5% females in the autistic group, to ensure sufficient sample size for meaningful conclusions about autistic females’ MH; (5) were COVID-19-related papers; (6) focused exclusively on a single pharmacological treatment; (7) were scale or tool validation studies; (8) addressed sleep problems without an explicit link to MH; (9) focused on eating specificities (e.g., food selectivity) without mentioning of eating disorders; or (10) were case studies, opinion pieces, editorials, or commentaries. Exceptionally, some of the latter (10) were retained if they offered insights into underexplored topics, for which larger-scale studies are limited (e.g., menstrual-related MH patterns, support strategies for MH in autistic females).

A data-charting form was developed by AL, MA, and M-CL, then refined after the first five extractions by the larger team. The final form included: study identification (first author name, publication year and title); continent (based on participants’ country of residence); aim; indication of whether the study was related to prevalence, support, maternity, female health, or biology; study design; sample ascertainment approach; type of autism diagnosis; age-period at autism diagnosis; number and age of participants in each group; type of MH issue reported; MH ascertainment; investigation of co-occurring ADHD, ID, other neurological or neurodevelopmental conditions, physical health issues, gender dysphoria, or genetic conditions; results about MH in autistic females; and biological findings. Data extraction was performed by one reviewer (AL, C-CT, BK-J, JXY, MA, or AW-B) and confirmed by AL.

To address objective 1 (i.e., mapping sex differences in the prevalence and presentation of MH conditions in autistic people), we employed two complementary approaches: an exploratory quantitative synthesis using meta-analytic approaches, and a narrative synthesis (also referred to as descriptive synthesis, King et al., 2022) for studies that could not be meta-analyzed. The narrative synthesis provides a structured summary of study findings, with attention to potential limitations (King et al., 2022). For the exploratory quantitative syntheses, we specifically identified population-based or registry-based (PopReg) studies that reported sex-disaggregated prevalence data. Given the rapid nature of this review, we did not conduct publication bias or quality assessments. Therefore, these exploratory syntheses should not be considered as formal meta-analyses. The quantitative syntheses were performed in R version 2025.05.1+513 (R Core Team, 2025) using the *metafor* package (Viechtbauer, 2010). For each included PopReg study, odds ratios (ORs) were calculated to compare the likelihood of each MH condition in autistic females versus males. ORs were log-transformed to normalize their distribution, stabilize variance, and ensure symmetrical confidence intervals (CIs). These log ORs were used as effect sizes in random-effects meta-analyses, applying restricted maximum likelihood (REML) estimation. Positive values indicate higher odds in autistic females; negative values indicate higher odds in autistic males. A log OR of 0 reflects no sex difference (OR = 1). The resulting pooled log ORs were subsequently back-transformed to obtain overall ORs for each MH condition. Between-study heterogeneity was assessed using the I² statistic and the Q-test for heterogeneity. Results of the meta-analytic models are reported with 95% CIs, p-values, I² values, and Q-test p-values for each MH condition. We restricted the meta-analytic syntheses to MH categories represented by at least four sex-stratified PopReg studies and with sufficient conceptual and measurement homogeneity (i.e., studies reporting clinical diagnoses, identified through ICD codes, medical records, or caregiver/self-report of a professional diagnosis). These included anxiety disorders, overall mood disorders, depressive disorders, bipolar disorders, ED, OCD, personality disorders, and schizophrenia spectrum and other psychotic disorders. Categories such as suicidality and SUD were excluded due to substantial heterogeneity in measurement and/or diagnostic definitions across studies (e.g., inconsistent inclusion of alcohol use disorder and other addictive behavioral disorders), but results of these categories were summarized in the narrative synthesis. Additional PopReg studies reporting sex differences without sex-stratified prevalence data, as well as clinical-based or community-based (ClinCom) studies and literature reviews, were also included in the narrative synthesis to complement the quantitative approach. Results of both (quantitative and narrative) syntheses are presented jointly by each MH condition, and discussed in relation to their reported sex-based patterns. We specified “children”, “adolescents”, “young adults”, “adults”, or “older adults”, depending on the age-group described (0–14, 10–19, 18–25, 19–65 and 65 years and older, respectively). These overlapping bands were chosen to (1) match with the age definitions reported in the primary studies, (2) align with standard definitions used by international organizations, and (3) preserve precision when summarizing findings from heterogeneous samples.

For objectives 2 to 4 (i.e., exploring modulating factors of MH, identifying explanatory mechanisms, and reporting support strategies), we summarized the findings using narrative synthesis only, given the diversity of methodologies in the included studies and the complexity or indirect nature of some associations with MH outcomes. Relevant studies were grouped into broad thematic categories to extract and organize key findings. No formal qualitative coding was performed.

## Results

### Search results

The database searches yielded a total of 10,415 articles and five additional records were identified through other sources. Then, 1,956 duplicates were removed automatically by Covidence and verified manually by AL. The team members (AL, C-CT, BK-J, JXY, MA, AW-B) removed 44 additional duplicates during abstract screening. Ultimately, 8,420 articles were screened, 2,334 full-text articles were assessed for eligibility, and 218 articles met the inclusion criteria (Figure 1). Among the included papers, 185 were original research articles (quantitative or qualitative), 12 were systematic reviews, scoping reviews, or meta-analyses, 11 were narrative reviews, 8 were case studies, and 2 were commentaries or viewpoints. Original research and case studies originated primarily from North America (n=86), followed by Europe (n=71), Australia (n=22), Asia (n=10), mixed continents (n=3), and Africa (n=1).

**Figure 1:**
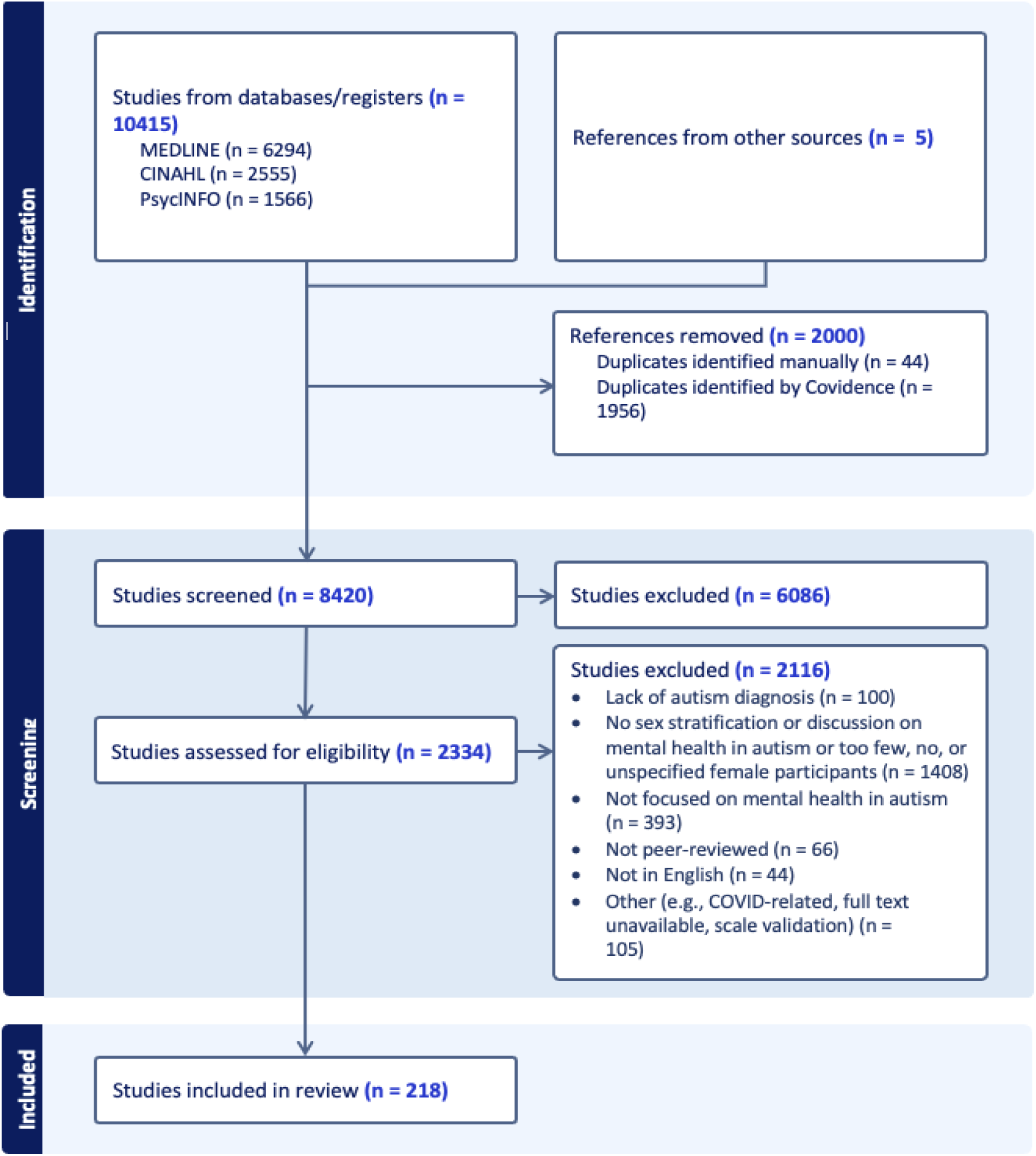
PRISMA flow diagram for study selection.

### MH prevalence and presentation in autistic females

Nineteen studies reported sex-stratified prevalence rates of MH conditions in autistic people or provided data allowing such calculations (Table 1). Of these, seventeen were suitable for inclusion in the exploratory meta-analytic syntheses, based on the criteria specified above (Figure 2, Table 2). These studies were complemented by an additional sixteen PopReg studies that, although not reporting sex-stratified prevalence rates, investigated sex differences in MH outcomes. Additional evidence came from 76 ClinCom studies, as well as three meta-analyses, one scoping review, one systematic review, six narrative reviews, and one commentary.

**Figure 2:**
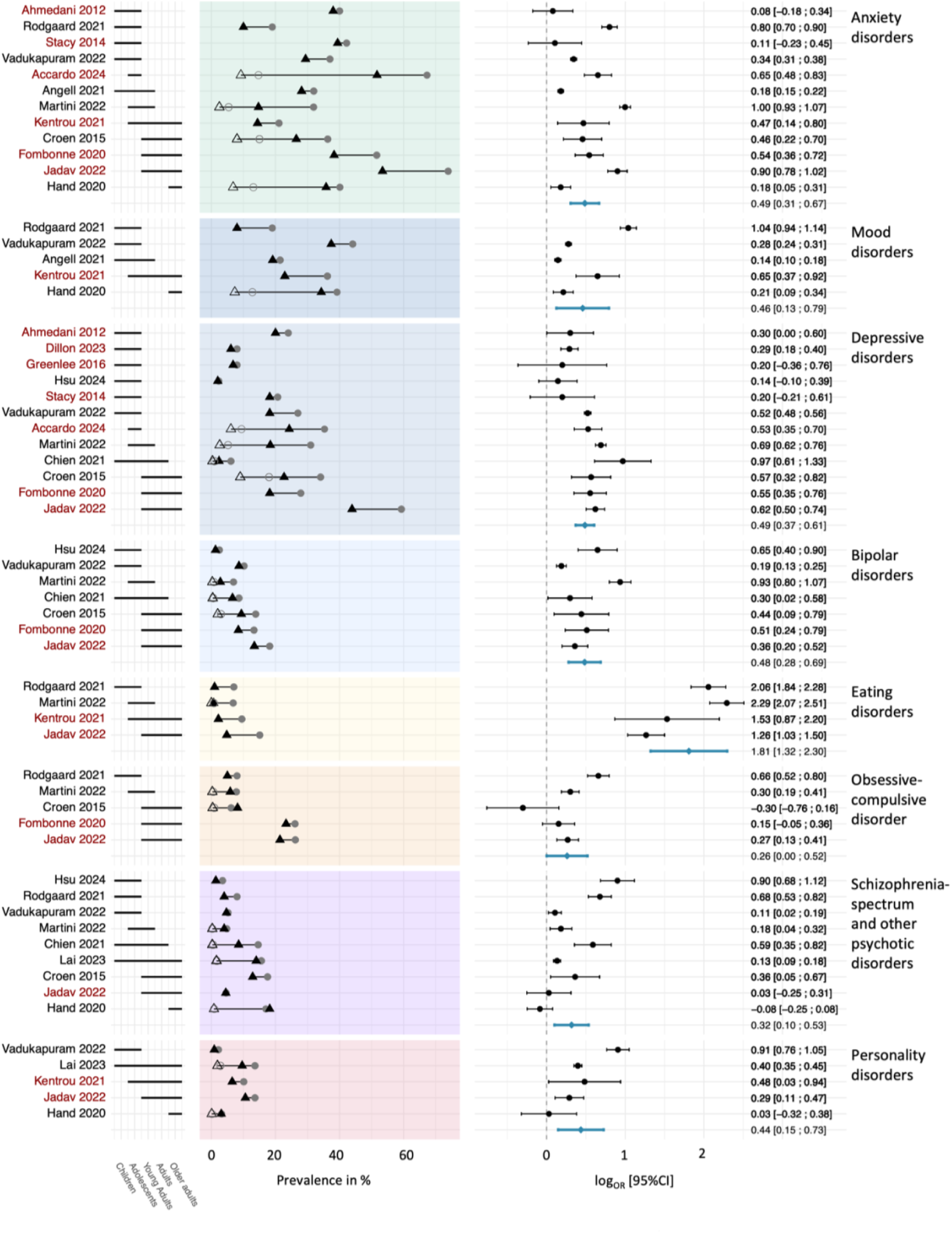
Observed prevalence in PopReg studies and exploratory meta-analytic synthesis. Left part: Observed prevalence of mental health (MH) conditions by study, ordered by age category, in autistic females (AF), autistic males (AM), non-autistic females (NAF), and non-autistic males (NAM). Study names in black correspond to studies with MH ascertainment based on ICD-codes; study names in dark-red correspond to studies with MH ascertainment based on self- or parental-report of clinical diagnosis. Age range (in year) covered by each category: children = 0-14, adolescents = 10-19, young adults = 18-25, adults = 19-65, and older adults = >65. Right part: Study-level (in black) and pooled meta-analytic estimates (in blue) of log odds ratios comparing the prevalence of MH conditions in autistic females versus autistic males, with 95% confidence intervals.

**Table 1:**
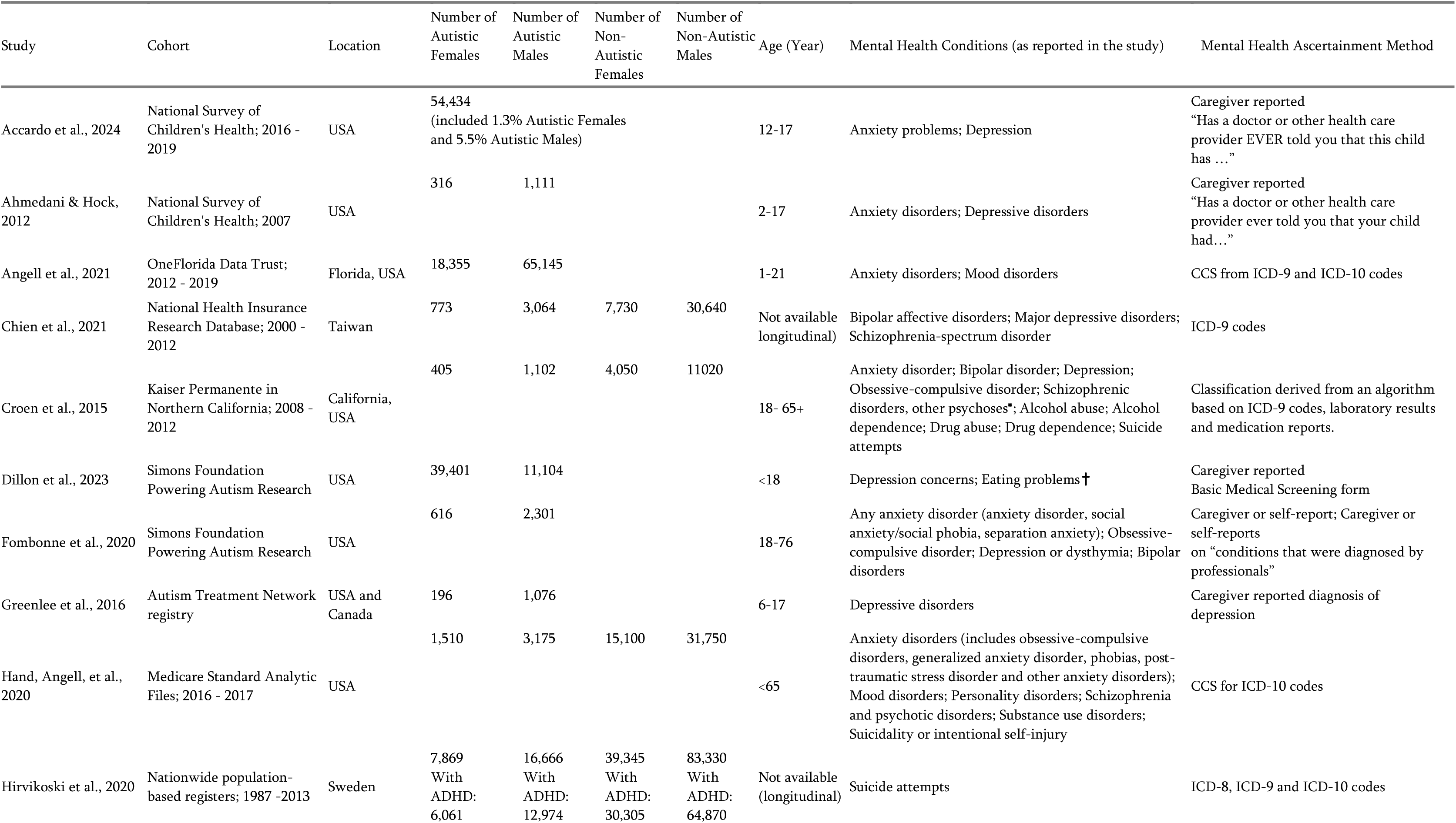
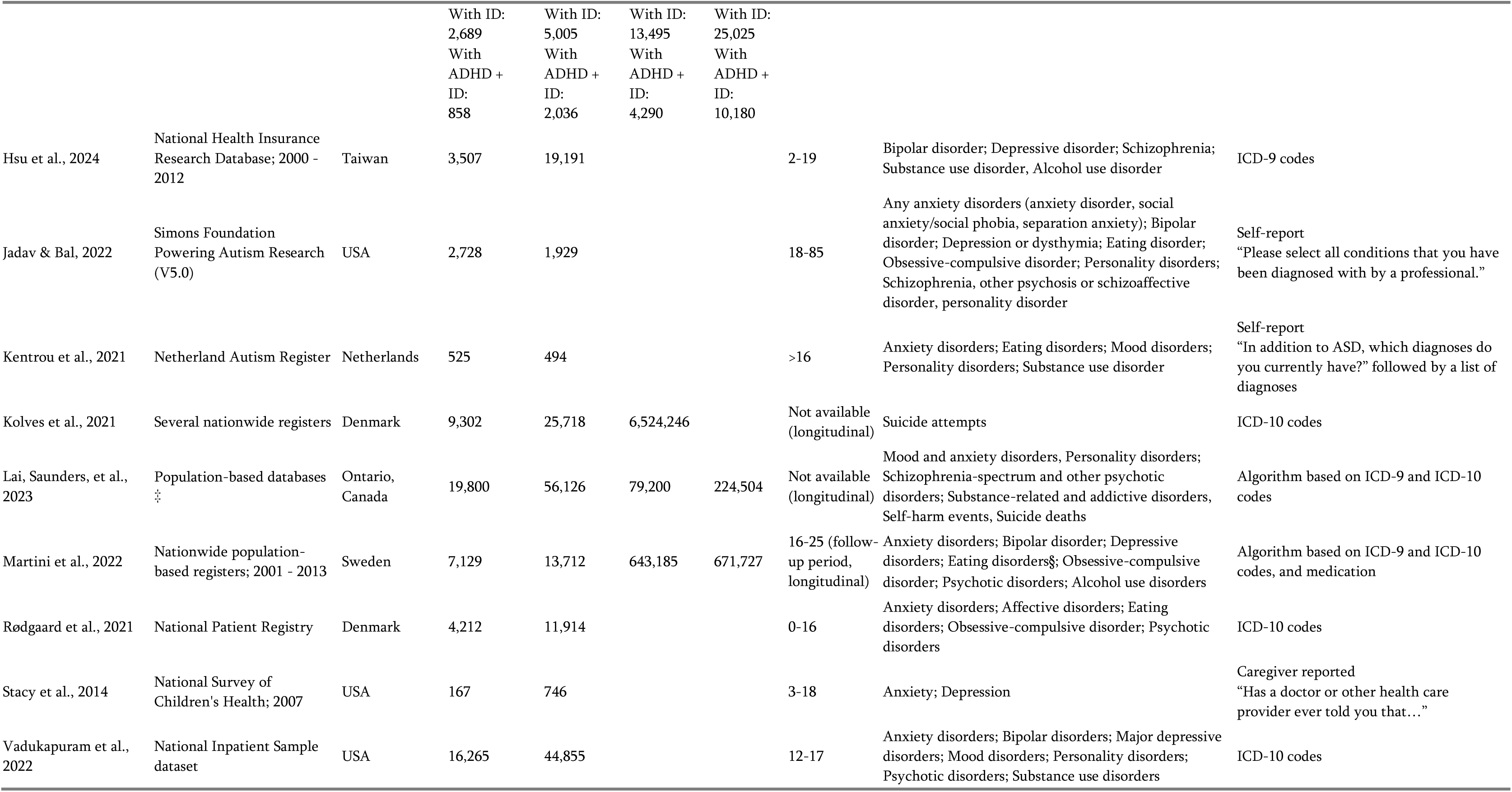
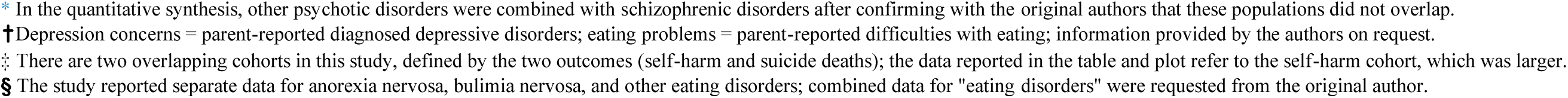
Characteristics of PopReg studies reporting sex-stratified data on mental health conditions in autistic people.

| Study | Cohort | Location | Number of Autistic Females | Number of Autistic Males | Number of Non-Autistic Females | Number of Non-Autistic Males | Age (Year) | Mental Health Conditions (as reported in the study) | Mental Health Ascertainment Method |
| --- | --- | --- | --- | --- | --- | --- | --- | --- | --- |
| Accardo et al., 2024 | National Survey of Children's Health; 2016 - 2019 | USA | 54,434<br>(included 1.3% Autistic Females and 5.5% Autistic Males) |  |  |  | 12-17 | Anxiety problems; Depression | Caregiver reported<br>"Has a doctor or other health care provider EVER told you that this child has ..." |
| Ahmedani & Hock, 2012 | National Survey of Children's Health; 2007 | USA | 316 | 1,111 |  |  | 2-17 | Anxiety disorders; Depressive disorders | Caregiver reported<br>"Has a doctor or other health care provider ever told you that your child had..." |
| Angell et al., 2021 | OneFlorida Data Trust; 2012 - 2019 | Florida, USA | 18,355 | 65,145 |  |  | 1-21 | Anxiety disorders; Mood disorders | CCS from ICD-9 and ICD-10 codes |
| Chien et al., 2021 | National Health Insurance Research Database; 2000 - 2012 | Taiwan | 773 | 3,064 | 7,730 | 30,640 | Not available (longitudinal) | Bipolar affective disorders; Major depressive disorders; Schizophrenia-spectrum disorder | ICD-9 codes |
| Croen et al., 2015 | Kaiser Permanente in Northern California; 2008 - 2012 | California, USA | 405 | 1,102 | 4,050 | 11020 | 18- 65+ | Anxiety disorder; Bipolar disorder; Depression; Obsessive-compulsive disorder; Schizophrenic disorders, other psychoses*; Alcohol abuse; Alcohol dependence; Drug abuse; Drug dependence; Suicide attempts | Classification derived from an algorithm based on ICD-9 codes, laboratory results and medication reports. |
| Dillon et al., 2023 | Simons Foundation Powering Autism Research | USA | 39,401 | 11,104 |  |  | <18 | Depression concerns; Eating problems† | Caregiver reported<br>Basic Medical Screening form |
| Fombonne et al., 2020 | Simons Foundation Powering Autism Research | USA | 616 | 2,301 |  |  | 18-76 | Any anxiety disorder (anxiety disorder, social anxiety/social phobia, separation anxiety); Obsessive-compulsive disorder; Depression or dysthymia; Bipolar disorders | Caregiver or self-report; Caregiver or self-reports on "conditions that were diagnosed by professionals" |
| Greenlee et al., 2016 | Autism Treatment Network registry | USA and Canada | 196 | 1,076 |  |  | 6-17 | Depressive disorders | Caregiver reported diagnosis of depression |
| Hand, Angell, et al., 2020 | Medicare Standard Analytic Files; 2016 - 2017 | USA | 1,510 | 3,175 | 15,100 | 31,750 | <65 | Anxiety disorders (includes obsessive-compulsive disorders, generalized anxiety disorder, phobias, post-traumatic stress disorder and other anxiety disorders); Mood disorders; Personality disorders; Schizophrenia and psychotic disorders; Substance use disorders; Suicidality or intentional self-injury | CCS for ICD-10 codes |
| Hirvikoski et al., 2020 | Nationwide population-based registers; 1987 -2013 | Sweden | 7,869<br>With<br>ADHD:<br>6,061 | 16,666<br>With<br>ADHD:<br>12,974 | 39,345<br>With<br>ADHD:<br>30,305 | 83,330<br>With<br>ADHD:<br>64,870 | Not available (longitudinal) | Suicide attempts | ICD-8, ICD-9 and ICD-10 codes |
|  |  |  | With ID: 2,689<br>With ADHD + ID: 858 | With ID: 5,005<br>With ADHD + ID: 2,036 | With ID: 13,495<br>With ADHD + ID: 4,290 | With ID: 25,025<br>With ADHD + ID: 10,180 |  |  |  |
| Hsu et al., 2024 | National Health Insurance Research Database; 2000 - 2012 | Taiwan | 3,507 | 19,191 |  |  | 2-19 | Bipolar disorder; Depressive disorder; Schizophrenia; Substance use disorder, Alcohol use disorder | ICD-9 codes |
| Jadav & Bal, 2022 | Simons Foundation Powering Autism Research (V5.0) | USA | 2,728 | 1,929 |  |  | 18-85 | Any anxiety disorders (anxiety disorder, social anxiety/social phobia, separation anxiety); Bipolar disorder; Depression or dysthymia; Eating disorder; Obsessive-compulsive disorder; Personality disorders; Schizophrenia, other psychosis or schizoaffective disorder, personality disorder | Self-report<br>“Please select all conditions that you have been diagnosed with by a professional.” |
| Kentrou et al., 2021 | Netherland Autism Register | Netherlands | 525 | 494 |  |  | >16 | Anxiety disorders; Eating disorders; Mood disorders; Personality disorders; Substance use disorder | Self-report<br>“In addition to ASD, which diagnoses do you currently have?” followed by a list of diagnoses |
| Kolves et al., 2021 | Several nationwide registers | Denmark | 9,302 | 25,718 | 6,524,246 |  | Not available (longitudinal) | Suicide attempts | ICD-10 codes |
| Lai, Saunders, et al., 2023 | Population-based databases | Ontario, Canada | 19,800 | 56,126 | 79,200 | 224,504 | Not available (longitudinal) | Mood and anxiety disorders, Personality disorders; Schizophrenia-spectrum and other psychotic disorders; Substance-related and addictive disorders, Self-harm events, Suicide deaths | Algorithm based on ICD-9 and ICD-10 codes |
| Martini et al., 2022 | Nationwide population-based registers; 2001 - 2013 | Sweden | 7,129 | 13,712 | 643,185 | 671,727 | 16-25 (follow-up period, longitudinal) | Anxiety disorders; Bipolar disorder; Depressive disorders; Eating disorders; Obsessive-compulsive disorder; Psychotic disorders; Alcohol use disorders | Algorithm based on ICD-9 and ICD-10 codes, and medication |
| Rødgaard et al., 2021 | National Patient Registry | Denmark | 4,212 | 11,914 |  |  | 0-16 | Anxiety disorders; Affective disorders; Eating disorders; Obsessive-compulsive disorder; Psychotic disorders | ICD-10 codes |
| Stacy et al., 2014 | National Survey of Children's Health; 2007 | USA | 167 | 746 |  |  | 3-18 | Anxiety; Depression | Caregiver reported<br>“Has a doctor or other health care provider ever told you that...” |
| Vadukapuram et al., 2022 | National Inpatient Sample dataset | USA | 16,265 | 44,855 |  |  | 12-17 | Anxiety disorders; Bipolar disorders; Major depressive disorders; Mood disorders; Personality disorders; Psychotic disorders; Substance use disorders | ICD-10 codes |

\* In the quantitative synthesis, other psychotic disorders were combined with schizophrenic disorders after confirming with the original authors that these populations did not overlap.
† Depression concerns = parent-reported diagnosed depressive disorders; eating problems = parent-reported difficulties with eating; information provided by the authors on request.
‡ There are two overlapping cohorts in this study, defined by the two outcomes (self-harm and suicide deaths); the data reported in the table and plot refer to the self-harm cohort, which was larger.
§ The study reported separate data for anorexia nervosa, bulimia nervosa, and other eating disorders; combined data for "eating disorders" were requested from the original author.

**Table 2:** Pooled prevalence, log odds ratios (OR) with 95% confidence intervals (CI), and OR comparing autistic females and males across mental health conditions, heterogeneity estimates (I², Q-test), and study sample sizes of autistic females (n F) and autistic males (n M).

| Mental Health<br>(N studies)<br>n Females<br>n Males | Pooled prevalence Autistic<br>females<br>[95% CI] | Pooled prevalence<br>Autistic males<br>[95% CI] | logOR [95%<br>CI] | p-value | Corresponding<br>OR | $I^2$, $Q$ test p-value |
| --- | --- | --- | --- | --- | --- | --- |
| Anxiety disorders<br>(N = 12)<br>n F = 52,937<br>n M = 149,478 | 40.4%<br>[31.4–50.2] | 29.7%<br>[22.1–38.6] | 0.49<br>[0.31–0.67] | <0.001 | 1.63 | 98.0%<br>($p \leq 0.001$) |
| Mood disorders<br>(N = 5)<br>n F = 40,867<br>n M = 125,583 | 31.1%<br>[22.8–40.8] | 22.1%<br>[13.3–34.4] | 0.46<br>[0.13–0.79] | 0.007 | 1.58 | 99.2%<br>($p \leq 0.001$) |
| Depressive disorders<br>(N = 12)<br>n F = 38,492<br>n M = 117,343 | 19.2%<br>[11.3–30.6] | 12.9%<br>[7.7–21.1] | 0.49<br>[0.37–0.61] | <0.001 | 1.63 | 88.0%<br>($p \leq 0.001$) |
| Bipolar disorders<br>(N = 7)<br>n F = 31,423<br>n M = 86,154 | 9.3%<br>[5.9–14.3] | 5.9%<br>[3.4–10.2] | 0.48<br>[0.28–0.69] | <0.001 | 1.62 | 90.0%<br>($p \leq 0.001$) |
| Eating disorders<br>(N = 4)<br>n F = 14,594<br>n M = 28,049 | 9.1%<br>[6.5–12.6] | 1.6%<br>[0.8–3.4] | 1.81<br>[1.32–2.30] | <0.001 | 6.10 | 92.2%<br>($p \leq 0.001$) |
| Obsessive-compulsive<br>disorder<br>(N = 5)<br>n F = 15,090<br>n M = 30,958 | 12.7%<br>[7.1–21.8] | 10.5%<br>[5.6–18.6] | 0.26<br>[0.00–0.52] | 0.049 | 1.30 | 91.7%<br>($p \leq 0.001$) |
| Schizophrenia-spectrum<br>and other psychotic<br>disorders<br>(N = 9)<br>n F = 56,329<br>n M = 155,068 | 8.5%<br>[5.6–12.7] | 6.3%<br>[3.8–10.3] | 0.32<br>[0.10–0.53] | 0.004 | 1.37 | 96.2%<br>($p \leq 0.001$) |
| Personality disorders<br>(N = 5)<br>n F = 40,828<br>n M = 106,579 | 6.8%<br>[3.4–13.1] | 4.5%<br>[2.0–9.9] | 0.44<br>[0.15–0.73] | 0.003 | 1.55 | 93.4%<br>($p \leq 0.001$) |

Most PopReg studies reported a higher prevalence of at least one psychiatric diagnosis in autistic females (36–77%) compared to autistic males (32–66%) (Kentrou et al., 2021; Martini et al., 2022; Rubenstein et al., 2018; Rydzewska et al., 2018), even after controlling for cognitive impairment, age, and language level (Fombonne et al., 2020). Similar patterns were observed in ClinCom studies (Baldwin & Costley, 2016; L. Bradley et al., 2021). This sex difference mirrors those observed in non-autistic populations (Martini et al., 2022; Rydzewska et al., 2018), but the extent was greater in autistic populations (Martini et al., 2022). Autistic adult females also had more psychiatric conditions than males (mean of 3 vs. 2.5; Jadav & Bal (2022); see also Dubreucq et al., 2023) and were more likely to have experienced psychiatric hospitalization by 25 years of age (Martini et al., 2022; 32% vs. 19%).

### Anxiety disorders

#### Anxiety disorders

Across twelve PopReg studies assessing anxiety disorders broadly, the pooled prevalence was 40.4% for autistic females and 29.7% for autistic males (OR = 1.63, *p* < 0.001), mirroring trends in the general and non-autistic populations (Croen et al., 2015; Hand, Angell, et al., 2020; Kirsch et al., 2020). Two additional PopReg studies did not report sex-disaggregated rates but examined the effect of sex on anxiety disorders, and found higher odds in autistic females than males in children, adolescents (Kerns et al., 2020) and adults (Kirsch et al., 2020). However, the increase in anxiety disorders compared to non-autistic people appeared more pronounced in autistic males than females (Croen et al., 2015; Hand, Angell, et al., 2020; Kirsch et al., 2020).

In contrast, except one study that found higher anxiety disorders in autistic adolescent females than males (So et al., 2021), most ClinCom studies did not report sex differences in the prevalence of anxiety disorders in autistic children, adolescents (Amr et al., 2012; Margari et al., 2019; Salazar et al., 2015), and adults (Lever & Geurts, 2016; Lugnegard et al., 2011; Tsakanikos et al., 2011). The absence of sex difference aligns with the summary from a systematic review (Bougeard et al., 2021), although only two of the included studies reported sex-stratified prevalence for anxiety disorders, limiting the strength of conclusions of this review. A narrative review also reported mixed results in sex differences in anxiety disorders in autistic children (Rubenstein et al., 2015).

Some studies further considered specific anxiety disorders. Social anxiety disorder (social phobia) and separation anxiety disorder were more prevalent in autistic adult females than males in two PopReg studies (Fombonne et al., 2020; Jadav & Bal, 2022). In contrast, no sex differences were reported in ClinCom studies of autistic children for the diagnoses of social phobia (Salazar et al., 2015) and separation anxiety disorder (Amr et al., 2012; Salazar et al., 2015), specific phobia (Amr et al., 2012; Salazar et al., 2015), panic disorder or agoraphobia (Salazar et al., 2015).

#### Anxiety symptoms

*Levels.* One PopReg study (Dubin et al., 2015) reported a higher likelihood of high anxiety symptoms in autistic male than female children and adolescents. In contrast, one ClinCom study indicated a higher likelihood for autistic female children to be above the cut-off of clinically significant anxiety problems than males, but this difference was not significant (Estes et al., 2023). Another ClinCom study in autistic adolescents and adults reported that significantly more females than males met the cut-off for clinically significant generalized anxiety disorder (Uljarevic et al., 2020).

Regarding anxiety intensity, one PopReg study reported no sex difference in generalized, social or separation anxiety symptoms (Jolly et al., 2020). ClinCom studies provided mixed results. Some reported higher anxiety symptoms in females than males in autistic children, adolescents (Long et al., 2024; Uljarevic et al., 2020), and adults (Anderson et al., 2020; Kuzminskaite et al., 2020; Uljarevic et al., 2020), but others found no sex differences, including those conducted with children and adolescents (May et al., 2014; May, Cornish, & Rinehart, 2015), adults (Lai et al., 2011, 2017; Lever & Geurts, 2016), or samples of all ages (Charman et al., 2017).

For symptoms of specific types of anxiety, one ClinCom study reported higher social anxiety symptoms in female children than in males, with no interaction with autism diagnosis, although social anxiety symptoms were higher in autistic than in non-autistic children (May et al., 2014). In the same study, autistic children also showed elevated symptoms of separation anxiety, panic, agoraphobia and generalized anxiety, with no effect of sex or interaction. In autistic female children, social anxiety may occasionally present as an unusually strong emotional attachment to a relative, as illustrated in a case study (Bhardwaj et al., 2005).

#### Anxiety symptoms

*Dimensions.* Some studies examined sex differences in specific dimensions of anxiety symptoms among autistic individuals. Ambrose et al. (2020) reported higher anxious arousal scores in autistic female children than in males, particularly for physiological symptoms, while performance anxiety and uncertainty subscales showed no sex differences.

Baldwin and Costley (2016) found no sex differences in self-reported experience of stress or worry among autistic adults, with 82% of female and 79% of male participants endorsing these experiences. In contrast, McGillivray and Evert (2018) reported that sex differences in stress levels varied by event type: compared to autistic males (aged 13 years and older), autistic females reported higher stress in response to pleasant events (e.g., socializing), sensory/personal contact (e.g., hugs, noise), and social/environmental interactions (e.g., asserting oneself, bright lights), while no significant difference was found for ritual-related stress, changes and social threats, unpleasant events, anticipation/uncertainty, and food-related activities. Additionally, autistic adult females (aged 25 and older) were especially sensitive to change, social threat, and uncertainty compared to autistic adult males, young males (13–24 years), and young females.

### Mood disorders

#### Mood disorders

Among five PopReg studies, the pooled prevalence of overall mood [affective] disorders (including major depressive disorder, dysthymia, and bipolar disorders) was 31.1% for autistic females and 22.1% for autistic males (OR = 1.58, *p* = 0.007). This pattern was consistent with a ClinCom study including autistic and non-autistic adults, showing that female sex significantly predicted the presence of any mood disorder (Lever & Geurts, 2016).

#### Depressive disorders

Twelve PopReg studies provided data on depressive disorders, leading to a pooled prevalence of 19.2% in autistic females and 12.9 % in autistic males (OR = 1.63, *p* < 0.001), mirroring the higher prevalence of depressive disorders in females than males in non-autistic populations (Chien et al., 2021; Croen et al., 2015; Martini et al., 2022). Importantly, variability in diagnostic labels (e.g., “depression”, “MDD”, “dysthymia”) and ascertainment methods across studies (Table 1) limits the precision of the pooled prevalence estimate. This result is nevertheless consistent with an additional PopReg study showing higher cumulative incidence of depression in autistic adult females than males (Kirsch et al., 2020). In contrast, another PopReg study in autistic children and adolescents found no sex difference (Kerns et al., 2020).

ClinCom studies reported fewer sex differences. One indicated that autistic adult females were more likely to have received a depression diagnosis than autistic males (Zheng et al., 2021). In contrast, five studies found no sex difference for depressive disorders in autistic children, adolescents (Margari et al., 2019) and adults (Tsakanikos et al., 2011), for major depressive disorder in children and adolescents (Amr et al., 2012; Salazar et al., 2015), for life-time/single episode/recurrent episode of major depression in adults (Lugnegard et al., 2011), and for dysthymia in children and adolescents (Albores-Gallo et al., 2017). Finally, a narrative review also reported mixed results about sex differences in depressive disorders in autistic children (Rubenstein et al., 2015).

PopReg studies showed elevated depressive disorders in autistic individuals compared to non-autistic populations (Accardo et al., 2024; Chien et al., 2021; Croen et al., 2015; Martini et al., 2022), suggesting an additive risk for females. However, the increased odds of depressive disorders in autistic versus non-autistic people was greater in males than in females (Croen et al., 2015; Kirsch et al., 2020).

#### Depressive symptoms

Regarding symptom intensity, one PopReg study showed higher depressive symptoms in autistic female children and adolescents than males (Long et al., 2024), whereas another found no sex differences (Jolly et al., 2020). ClinCom studies reported mixed results. Some showed greater depressive symptom severity in autistic females than males (Anderson et al., 2020; Burns et al., 2019; Charman et al., 2017; Corbett, Muscatello, et al., 2024; Costa et al., 2020; Hedley et al., 2018; McGillivray & Evert, 2018; Schwartzman et al., 2022; Smith et al., 2024; Uljarevic et al., 2020) and one ClinCom study reported that autistic adolescent and adult females were more likely than males to score above the clinical cut-off for depression (Uljarevic et al., 2020). As autistic individuals generally experienced higher rates of depressive symptoms compared to non-autistic individuals (Bitsika, Sharpley, & Mills, 2021; Corbett, Muscatello, et al., 2024; Martini et al., 2022; Schwartzman et al., 2022), Schwartzman et al. (2022) suggested an additive risk for autistic females. Nevertheless, other studies found no sex difference in symptom severity (Holtmann et al., 2007; Lai et al., 2011, 2017; Schwartzman & Bonner, 2024; Zheng et al., 2021).

#### Bipolar disorders

From the seven PopReg studies that provided sex-disaggregated data for bipolar disorders, the pooled prevalence was 9.3% for autistic females and 5.9% for autistic males (OR = 1.62, *p* < 0.001). This result is consistent with a meta-analysis showing an increased likelihood of bipolar disorders in autistic females compared to males (Varcin et al., 2022). In contrast, a ClinCom study found no sex difference (Margari et al., 2019). Notably, one PopReg study also found that the odds of bipolar disorders were similarly elevated in autistic females and males compared with their non-autistic counterparts (Croen et al., 2015).

### Eating Disorders

#### Eating disorder (ED)

A significant sex difference emerged from the four PopReg studies related to ED, with autistic females showing a pooled prevalence of 9.1% compared to 1.6% in autistic males (OR = 6.10, *p* < 0.001), a pattern also seen in non-autistic individuals (Martini et al., 2022). However, the relative risk compared to non-autistic individuals was greater in males (Martini et al., 2022). In contrast, another PopReg study (Dillon et al., 2023) found a higher prevalence in autistic males than females, potentially reflecting a broader definition of “eating problems”, referring to problems eating food rather than clinically diagnosed eating disorder, which may explain the high overall prevalence (38% of autistic females and 42% of autistic males). ClinCom findings were mixed: one study reported more anorexia nervosa in autistic female children and adolescents compared to autistic males (Margari et al., 2019), while two found no sex difference for ED in autistic adults (Karjalainen et al., 2016; Lugnegard et al., 2011).

#### Symptoms of disordered eating

Adolescent females with Asperger syndrome, compared to those without, reported more eating-related problems, especially bulimia, food preoccupation and oral control, but not more dieting behaviors; a greater proportion exceeded the cut-off for referral for ED assessment (Kalyva, 2009). Emotional over- and under-eating were also more frequent in autistic children and adolescents, with over-eating more often reported by females than males, a sex difference not seen in the non-autistic sample (Wallace et al., 2021).

### Obsessive-compulsive disorder (OCD)

The prevalence of OCD among autistic females was estimated at 12.7%, compared to 10.5% in males (OR = 1.30, *p* = 0.049), across five PopReg studies. However, the odds of having OCD compared to non-autistic people were higher in autistic males than females (Croen et al., 2015; Martini et al., 2022). Importantly, the prevalence of OCD in autistic people varied widely depending on ascertainment, ranging from 5–8% when identified through International Classification of Diseases (ICD) codes (Croen et al., 2015; Martini et al., 2022; Rødgaard et al., 2021), to 21–42 % when based on self- or caregiver reports (Fombonne et al., 2020; Jadav & Bal, 2022).

A ClinCom study showed no sex differences for OCD in autistic children (Amr et al., 2012). Another suggested that autistic female children with OCD presenting with anxiety symptoms might be particularly prone to hoarding behaviors compared to males (La Buissonniere-Ariza et al., 2018). While this may indicate sex differences in OCD symptom profile in autistic people, further evidence is required.

### Schizophrenia spectrum and other psychotic disorders

The pooled prevalence of schizophrenia spectrum and other psychotic disorders among autistic individuals was estimated at 8.5% for females and 6.3% for males across nine PopReg studies (OR = 1.37, *p* = 0.004). Among these, one study reported prevalence for schizophrenia only (Hsu et al., 2024), and was still included in the pooled estimates, which may have slightly underestimated the true prevalence of all psychotic disorders. When analyzed, odds of schizophrenia and other psychotic disorders in autistic compared to non-autistic individuals were greater in females than males (Croen et al., 2015; Martini et al., 2022).

However, a meta-analysis including seven studies found a higher likelihood of psychosis in autistic adult males than females (Varcin et al., 2022), and a ClinCom study reported more schizophrenia in autistic adult males than females (Tsakanikos et al., 2011). In addition, a PopReg study in adults showed a significant association between autism and schizophrenia in males only (Krieger et al., 2021) and a ClinCom study in autistic adults with or without psychosis reported fewer females in the autism-with-psychosis group (Larson et al., 2015). Meanwhile, a ClinCom study found no sex difference in autistic adults for psychotic disorders (Lugnegard et al., 2011). Finally, one study on psychotic experiences found more lifetime subclinical symptoms of psychosis phenotype, comparable lifetime psychotic experiences, but greater associated distress in autistic than non-autistic adults, with no sex difference (van der Linden et al., 2020).

### Personality disorders

Among the five PopReg studies reporting sex-stratified data for personality disorders, the pooled prevalence was 6.8% for autistic females and 4.5% for autistic males (OR = 1.55, *p* = 0.003). Rates were higher than those in the non-autistic populations for both sexes (Hand, Angell, et al., 2020; Lai, Saunders, et al., 2023). In contrast, older ClinCom studies showed higher rates in autistic adult males than females (Lugnegård et al., 2012; Tsakanikos et al., 2011), all belonging to clusters A (schizotypal) and C (obsessive-compulsive, avoidant) (Lugnegård et al., 2012). Nevertheless, a ClinCom study of adults referred for borderline personality disorder (cluster B) found that among the 41 females diagnosed, 19 were assessed for suspected autism and six met the criteria for an autism diagnosis. These autistic females with borderline personality disorder had more suicide attempts, less substance abuse, lower self-love and self-control, and poorer global functioning than their non-autistic counterparts diagnosed with borderline personality disorder (Ryden et al., 2008).

### Suicidality

Five PopReg studies reported sex-stratified rates for suicide attempts or self-harm requiring emergency care, ranging 0.7–14% in autistic females and 0.15–6 % in autistic males (Croen et al., 2015; Hand, Benevides, et al., 2020; Hirvikoski et al., 2020; Kolves et al., 2021; Lai, Saunders, et al., 2023). Four studies reported higher rates in autistic females than males, although sex differences were not statistically analyzed (Croen et al., 2015; Hirvikoski et al., 2020; Kolves et al., 2021; Lai, Saunders, et al., 2023). A meta-analysis covering ten studies and over ten million individuals across seven countries found a greater risk of suicide attempt in autistic females compared to males (RR 3.85; Santomauro et al., 2024). All five PopReg studies also included non-autistic comparison groups, with consistently lower rates observed, i.e., 0.07–2.6% in non-autistic females and 0.05–2% in non-autistic males (Croen et al., 2015; Hand, Benevides, et al., 2020; Hirvikoski et al., 2020; Kolves et al., 2021; Lai, Saunders, et al., 2023). Studies on suicidality also broadly encompassed suicidal ideation and deaths, and analyzed different suicidal methods. Findings from large-scale ClinCom studies on suicidal ideation in autistic children and adolescents were mixed; one study identified male sex as a predictor (Conner et al., 2020) while two other reported higher risks in females than males (Anderson et al., 2020; Holden et al., 2020). Additionally, autistic adolescent females referred to emergency psychiatric services were found to be at greater risks of suicide and self-harm than males (So et al., 2021). However, most studies did not identify sex as a significant correlate of suicidal ideation (PopReg: Bentum et al., 2024; Hand, Benevides, et al., 2020; ClinCom: Arwert & Sizoo, 2020; Cassidy et al., 2018; Costa et al., 2020; Hedley et al., 2018; Hunsche et al., 2020; McDonnell et al., 2020; Pelton et al., 2023; Storch et al., 2013), consistent with the findings of a scoping review (Cleary et al., 2023).

Suicide deaths are typically examined in PopReg studies, with heterogeneous findings. One study reported higher suicide mortality in autistic males than females (although autistic females were more likely to die by suicide following severe self-harm; Hull et al., 2024), consistent with sex differences noted in the non-autistic population (Kirby 2019). However, other studies (Kirby et al., 2019; Kolves et al., 2021), a PopReg-focused commentary (Kirby et al., 2024) and a scoping review (Cleary et al., 2023) all concluded similar rates across sexes in the autistic population. Findings also varied regarding comparisons with non-autistic populations. Some studies suggested higher rates of suicide deaths in autistic compared non-autistic people (Hull et al., 2024; Tsai et al., 2023). Others reported more nuanced, sex-specific patterns. One study noted this trend only after 2013 and especially among autistic females (Kirby et al., 2019). Other studies found higher rates in autistic than non-autistic females, with no significant difference in males (Lai, Saunders, et al., 2023), or, conversely, greater risks in autistic than non-autistic males, with no significant difference in females (Tsai et al., 2023).

Regarding methods, non-violent means (e.g., poisoning) were most commonly used for severe self-harm among autistic individuals (Hull et al., 2024), whereas suicide deaths more often involved violent methods (e.g., firearms, hanging, blunt force; Kirby et al., 2019). These patterns were similar to those noted in the non-autistic population, although autistic people were less likely to use firearms (Kirby et al., 2019). This study also found no sex difference in suicide methods in autistic people, while a ClinCom study in an emergency department context reported more severe methods used by males, both in autistic and non-autistic groups (Mikami et al., 2019).

### Substance use disorders

#### Substance use disorders (SUD)

Five PopReg studies provided sex-stratified data related to SUD (Hsu et al., 2024; Kentrou et al., 2021; Lai, Saunders, et al., 2023; Martini et al., 2022; Vadukapuram et al., 2022). Four studies investigated SUD broadly (Hand, Angell, et al., 2020; Kentrou et al., 2021; Vadukapuram et al., 2022), with one including also other addictive disorders (Lai, Saunders, et al., 2023), one focused on alcohol use disorder (Martini et al., 2022) and another distinguished substance from alcohol use disorder (Hsu et al., 2024). Prevalence ranged 0.2–8.6% in autistic females and 0.2–10% in autistic males. One study found higher SUD rates in autistic adolescent females compared to males (Vadukapuram et al., 2022), while two reported no significant sex differences for alcohol use disorder (Hsu et al., 2024; Martini et al., 2022) or SUD (Hsu et al., 2024). In contrast, Croen et al. (2015) reported higher rates of alcohol abuse, drug abuse and drug dependence, but not alcohol dependence, in autistic adult males than females. Additionally, while two PopReg studies reported increased odds of SUD and other addictive disorders (Lai, Saunders, et al., 2023) and alcohol use disorder (Martini et al., 2022), in both autistic females and males compared to non-autistic individuals, others did not find elevated rates of SUD (Hand, Angell, et al., 2020) or alcohol/drug abuse or dependence (Croen et al., 2015) in autistic people. The absence of sex difference in SUD was also found in a ClinCom study (Lugnegard et al., 2011).

#### Substance use behaviors

A large-scale study (N autism = 1,183) found that autistic individuals were less likely than non-autistic individuals to consume alcohol regularly or to engage in binge drinking, and autistic males were less likely than non-autistic males to have ever smoked or used drugs, a difference not observed in females (Weir et al., 2021). Finally, a ClinCom study reported higher alcohol use scores in autistic males compared to females (Bowri et al., 2021).

### Factors modulating MH in autistic females

The first part of this review suggests that autistic females are at increased risks for MH difficulties. Several factors might modulate their MH outcomes differently compared to autistic males. Based on the current literature, we identified nine individual-level factors that might especially influence MH in autistic females.

#### Sex-related physiology

The influences of sex-specific and reproductive health factors include female puberty, menstruation, pregnancy, and menopause. Families of autistic females were more likely to express concerns about anticipated behavioral changes during puberty compared to families of females with other special needs, as puberty could exacerbate pre-existing challenges such as sensory sensitivities and emotion dysregulation (Fei et al., 2021). These concerns were supported by findings that autistic children and adolescents who reached menarche were rated by their mothers as more worried, irritable, tense, fatigued, and depressed than those who had not (Bitsika, Sharpley, & Mills, 2021; Bitsika & Sharpley, 2020). However, menarche status explained only a small part of the variance in depression ratings, whether self-reported or reported by mothers, suggesting that hormonal changes alone did not fully explain the increase in depression with age, and that psychosocial and environmental factors might interact with biological processes to exacerbate symptoms (Sharpley et al., 2021).

The menstrual cycle was also described by professionals as challenging for autistic females, sometimes leading to increased anxiety and self-injurious behaviors (Lundin et al., 2021). Autistic adult females without ID exhibited higher prevalence (14–21%) of *Premenstrual Dysphoric Disorder* (PMDD) than non-autistic females (3–9%) (Groenman et al., 2022; Lever & Geurts, 2016). Among the studies reviewed, only one investigated PMDD in autistic adult females with ID (Obaydi & Puri, 2008). Caregiver and nurse ratings of behavior and mood over three consecutive menstrual cycles revealed that 92% of autistic females, compared to 11% of their non-autistic peers, fulfilled the DSM-IV criteria for late luteal phase dysphoric disorder (corresponding to DSM-5 PMDD) during the nine days preceding menstruation. All autistic participants experienced affective lability, anger or irritability, anxiety, and depressed mood. Additional symptoms included tantrums, destructive behavior, self-harm, physical aggression, as well as changes in sleep, autistic traits, appetite, and headaches. Further insight came from three case studies describing MH difficulties associated with premenstrual syndrome or menstruation in autistic females with ID. The first reported two autistic females who showed a drastic increase in mood symptoms, irritability, agitation, tantrums, aggressive behaviors, and amplified autistic behaviors (Lee, 2004). The authors emphasized the importance of recognizing PMDD as a contributing factor to mental and behavioral challenges to avoid inappropriate interventions. This was illustrated by a second case study describing an autistic female with bipolar disorder (Vijapura et al., 2014). At puberty, she developed a pattern of worsened irritability and increased manic symptoms three to four days before menstruation, typically resolving in the initial days of menstruation. At age 15, she experienced an episode of acute mania with hospitalization. Treatment with quetiapine and lamotrigine provided some relief, but was discontinued due to excessive sedation. A trial of oral contraceptives alone resulted in a year-long remission of manic symptoms, although symptoms later re-emerged, likely due to psychological stressors requiring additional treatment. This case underscores the pressing need for research on hormonal fluctuations and their impact on MH in autistic females, including those with ID. A third case study described severe depression, suicidal thoughts, and self-injurious behaviors triggered cyclically by menstruation in a young autistic female, alongside increased obsessive-compulsive behaviors related to hygiene and changes during this period (Skinner et al., 2005). Unfortunately, hormonal treatment had little success in this case, even leading to the consideration of definitive surgery, which posed important ethical challenges regarding the sterilization of individuals unable to give their own consent.

Autistic females may also become parents. Notably, pregnancy and parenthood may occur prior to a formal diagnosis of autism (Pohl et al., 2020). The perinatal period is therefore an important window for monitoring and support, as the related biopsychosocial changes can trigger MH challenges. Autistic females reported higher rates of pregnancy-related complications than non-autistic females such as pelvic pain and vaginal bleeding, which may partly contribute to their elevated likelihood of experiencing anxiety and depression during this period (Hampton et al., 2022, 2023). According to a qualitative study, childbirth may also be particularly distressing due to challenges in communicating needs, pain. and uncertainty caused by a lack of clear explanations from healthcare providers, and overstimulating environments during labor and postpartum. These factors can heighten anxiety and trigger shutdowns or withdrawal, which in turn might further impair communication and increase distress (Donovan, 2020). In line with this, autistic mothers were more likely to have greater stress, anxiety, and depression than non-autistic peers at each timepoint of the perinatal period (prenatal, 2–3 months post-partum, and 6 months post-partum) according to an in-person longitudinal study (Hampton et al., 2022). Meanwhile, a retrospective study (assessing autistic females up to 15 years after pregnancy) found no significant difference in postpartum depression (Demartini et al., 2024). The difference between the two studies might be partly explained by a recall bias. In the latter, however, autistic traits were associated with higher levels of postpartum depressive symptoms in the autistic but not the non-autistic group (Demartini et al., 2024). It remains unclear whether perinatal MH issues of autistic mothers are part of a broader vulnerability, as autistic mothers were also more likely to have received prior diagnoses of anxiety and depression than their non-autistic peers (Hampton et al., 2022). It might also reflect context-specific difficulties during this life stage. Regarding parenthood more broadly, autistic mothers reported levels of parenting stress similar to those of non-autistic mothers, despite higher rates of MH diagnoses (D. Adams et al., 2021).

Menopause is another major milestone in female reproductive health, with potential implications for both physical and mental well-being. A PopReg study showed that 4% of autistic females aged 35–70 years had symptomatic menopause (defined as seeking care or treatment and identified by medical claims), compared to 8% in the general population (Benevides et al., 2024). Another study, however, found no difference in prevalence (Hand, Angell, et al., 2020). While these findings seem to suggest a lower risk of symptomatic menopause in autistic people, they may also reflect barriers to recognizing and expressing menopausal difficulties among autistic females, especially those with ID, who had less symptomatic menopause claims than those without ID (Benevides et al., 2024). By contrast, in a sample composed exclusively of females without ID (aged 42 and above) and in which menopausal complaints were assessed by a questionnaire, psychological and somatic complaints were more frequent in autistic than in non-autistic females (Groenman et al., 2022). Psychological menopausal complaints were associated with depressive symptoms in both autistic and non-autistic groups, and with autistic traits specifically in the autistic group (Groenman et al., 2022). Similarly, a PopReg study found that autistic females with symptomatic menopause were more likely than autistic females with non-symptomatic menopause to report MH problems, such as anxiety and depression, as well as sexual dysfunction, migraine, altered sensory experiences, and sleep disturbances (Benevides et al., 2024).

Overall, these findings align with a narrative review emphasizing increased difficulties, particularly emotional ones, experienced by autistic females during puberty and menopause (Rynkiewicz et al., 2024). Our results also highlight a crucial gap in the literature regarding how sex-specific biological processes interact with autism to influence MH trajectories. Moreover, they underscore the longstanding lack of consideration and methodological challenges in assessing the impact of female-specific and reproductive health factors, particularly among autistic females with ID and/or limited language.

#### Gender-related experiences

Although several studies stratified populations based on either sex recorded at birth or self-identified gender, few combined both dimensions, and sample sizes of gender-diverse individuals were often limited. Nonetheless, existing evidence suggests that gender identity is a meaningful modulating variable of MH in autistic AFAB individuals. This relevance stems from two synergistic observations. First, gender-diverse or gender non-conforming autistic people are at elevated risk of MH difficulties, including anxiety, depression, self-harm, and suicidal ideation (Anderson et al., 2020); second, gender diversity is more frequently reported among autistic people than in the non-autistic population, and may be especially salient for autistic AFAB people (for a systematic review, see Bouzy et al., 2023). Several hypotheses have been proposed to explain the association between gender diversity and autism, involving social factors (e.g., heightened non-conformity with social norms) and biological mechanisms (e.g., influence of fetal testosterone or endocrine specificities). According to Bouzy et al. (2023), the sex ratio in co-occurring autism and transgender identity is either balanced (in six studies) or skewed toward AFAB (in six others). Puberty may contribute to this AFAB-predominance, as it involves more pronounced changes in AFAB than in AMAB individuals, which may provoke significant distress and contribute to the emergence of gender incongruence or dysphoria (Brunissen et al., 2021). Indeed, in the latter study, 81% of autistic AFAB were reported by their parents to experience discomfort during puberty, compared to 39% of autistic AMAB. Similarly, a greater proportion of parents of autistic AFAB (34%) than AMAB (15%) perceived that their child experienced anxiety related to gender concerns.

A clinical case study further illustrated the complexity of gender exploration in autistic AFAB and the importance of considering MH (Zupanic et al., 2021). It reported on an autistic AFAB adolescent who experienced co-occurring gender dysphoria, depression, and suicidality. The care team opted for a supportive, exploratory psychotherapeutic approach, prioritizing stabilization of MH before gender-affirming medical treatment. Over time, as the individual’s MH improved, gender dysphoria persisted and led to the initiation of hormonal treatment and the planning of a mastectomy. The authors emphasized that the process of gender exploration and potential transition in autistic individuals may require more time and careful MH support, and that a flexible, individualized approach is essential.

In sum, gender-diverse autistic people appear to be particularly vulnerable to MH difficulties, and gender-related concerns may represent a significant source of distress, especially among autistic AFAB people. These findings underscore the importance of integrating both sex- and gender-based perspectives into research on MH in autistic people, an area that still requires substantial empirical and clinical development.

#### Age

MH diagnostic patterns vary with age. For example (and by definition), Oppositional Defiant Disorders were more frequently reported in autistic children and adolescents, whereas personality disorders were more common in adults (Jadav & Bal, 2022). Interestingly, both the youngest and oldest autistic individuals were less likely to have psychiatric diagnoses overall (Jadav, 2022; Lever, 2016), suggesting MH difficulties may peak (or be most noticeable) during adolescence and early adulthood. Supporting this, older age was a significant predictor of all MH conditions in autistic children and adolescents (Kerns et al., 2020), and of suicidal behavior in autistic adolescents and adults (McDonnell et al., 2020; Santomauro et al., 2024). A longitudinal study described an increase in anxious and depressive symptoms from adolescence to middle adulthood, followed by a decline (Uljarevic et al., 2020). In contrast, another study observed a general reduction in internalizing and externalizing symptoms across a broad age range (10 to 48 years), without a clear peak period (Woodman et al., 2016).

Importantly, findings pointed to sex-specific trajectories. For example, autistic females in early childhood showed a faster reduction in externalizing symptoms compared to males (Wright et al., 2023), yet they were more likely to experience persistent internalizing symptoms over time (Vaillancourt et al., 2017). This is consistent with increased self- and parent-reported anxiety symptoms in autistic females between ages 6 and 15 years, while anxiety symptoms decreased among autistic males in the same period (Bitsika & Sharpley, 2020; Horwitz et al., 2023). Notably, similar patterns were reported in the non-autistic population (Horwitz et al., 2023). Whereas this suggests differences that are not unique to autism, the mechanisms underlying these different developmental trajectories across sex remain poorly understood. One study identified different correlates of self-reported anxiety across development in autistic females: maternal anxiety in childhood and physiological stress during adolescence as measured by diurnal cortisol levels (Bitsika, Sharpley, Mandy, et al., 2021). This implies certain developmental mechanisms underlying anxiety in autistic females that might differ in autistic males. In terms of depression, sex-specific patterns were similar. A PopReg study reported that before age 15, autistic males had a higher cumulative incidence of depression than females, whereas the pattern reversed after 15 with higher rates in autistic females (Kirsch et al., 2020). This developmental shift is supported by another study (Murray et al., 2019) that observed an earlier age-related decline in depression symptoms in autistic adult males compared to females, although the relationship between age and depression did not reach statistical significance.

In contrast, several studies did not find significant variations in MH trajectories by sex. For instance, anxiety disorders (Salazar et al., 2015) and symptoms (Mayes et al., 2011; but Bitsika, Sharpley, Mandy, et al., 2021) increased with age in autistic children, but none of these studies reported an interaction with sex. Similarly, stress levels increased between adolescence and adulthood, again with no interaction with sex (McGillivray & Evert, 2018). Depressive symptoms rose from childhood to adolescence (Greenlee et al., 2016; Mayes et al., 2011) and from adolescence to adulthood (McGillivray & Evert, 2018), with no sex-related differences in trajectories.

#### Age at autism diagnosis

Several studies showed higher rates of MH service contact (Dubreucq et al., 2023) and increased psychiatric diagnoses or symptoms, particularly anxiety, depression, and ED, prior to autism diagnosis in autistic female children, adolescents (Gu et al., 2023), and adults (Dubreucq et al., 2023; Geurts & Jansen, 2011; Kentrou et al., 2021; Murray et al., 2019) compared to their male peers. This was consistent with systematic review findings (Lockwood Estrin et al., 2021). Co-occurring diagnoses of anxiety, depression, and ADHD (but not ID) were associated with later autism diagnoses in female children and adolescents (Gu et al., 2023; Wodka et al., 2022) while they accelerated diagnosis in males (Gu et al., 2023). Accordingly, autistic females were more likely to be referred later for an autism assessment (Geurts & Jansen, 2011)) and diagnosed later than males (Gu et al., 2023; Kentrou et al., 2021; Smith et al., 2024). Autistic females were also more at risk for misdiagnosis. In one study of autistic adults, 63% of females and 37% of males reported at least one prior diagnosis, and more females than males (47% vs. 27%) reported that at least one, most often a personality disorder, was no longer considered a true co-occurring diagnosis (Kentrou et al., 2021).

Autism diagnosis delays can exacerbate MH difficulties, including psychological distress, anxiety, depression, and self-harm (Blainey et al., 2017; Fowler & O’Connor, 2021; Jadav & Bal, 2022; Rødgaard et al., 2021; Smith et al., 2024), as well as lifetime prevalence of psychiatric diagnoses (Jadav & Bal, 2022). Autistic females were particularly at risk. One study explored this pathway and found an indirect effect of sex on parent-reported anxious and depressive symptoms through age at autism diagnosis, such that females diagnosed later reported greater symptoms (Smith et al., 2024). Suicidal ideation (Fowler & O’Connor, 2021) and attempts (Kolves et al., 2021) were also more frequent among females diagnosed later. This exacerbation of MH difficulties in late-diagnosed individuals may be related to cumulative lifetime stress (Green et al., 2019). It may also stem from the distress associated with lacking a coherent framework to understand one’s differences (Blainey et al., 2017). In turn, this may hinder the development of self-identity and the integration of an autistic identity. In particular, camouflaging behaviors, often developed and sustained over many years before an autism diagnosis, can blur self-identity by contributing to internal conflict, and are associated with poorer MH (Halsall et al., 2021; see also Social Contributions section).

#### Autistic characteristics

Autistic characteristics represent another factor that may predispose autistic individuals, particularly females, to MH difficulties and may also serve as a moderating factor. One salient example is the development, maintenance, and treatment of ED (Schroder et al., 2023). In autistic people, traditional disordered eating symptoms appear closely intertwined with autism-specific features (Brede et al., 2020, 2024), leading to a modified clinical profile. This profile includes heightened sensory sensitivities (e.g., during mealtime and shopping), social difficulties during meals, emotional dysregulation, restricted and repetitive behaviors, and rigid thinking (Bitsika & Sharpley, 2018; Brede et al., 2024; C. M. Brown & Stokes, 2020; Schroder et al., 2023; Sharpley & Bitsika, 2022). These traits were elevated in autistic females with ED compared to both autistic females without ED and non-autistic females with ED (Brede et al., 2024). They may contribute to eating difficulties (Brede et al., 2020, 2024; Sharpley & Bitsika, 2022; see also the review of C. M. Brown & Stokes, 2020), which sometimes serve as maladaptive coping strategies (Brede et al., 2020; Schroder et al., 2023). Restricted and repetitive behaviors (RRBs) in particular may lead to hyperfocus on health, dieting, or exercise routines (Schroder et al., 2022). This may partly explain why restrictive ED in autistic females began earlier and were diagnosed sooner than in non-autistic females (Babb et al., 2022), often presenting more severely at treatment onset (Schroder et al., 2023; Zhang et al., 2022). Treatment duration might also be longer in this population (Babb et al., 2022; Zhang et al., 2022; and scoping review of Beygui & Cascio, 2022) with more frequent use of tube feeding (Zhang et al., 2022) and poorer outcomes, likely due to interventions not tailored to autistic characteristics (Beygui & Cascio, 2022; Schroder et al., 2022). In contrast, some ED symptoms appeared less prominent in autistic females than in their non-autistic peers: lower levels of body image dissatisfaction, fewer concerns related to weight and shape (Babb et al., 2022; Brede et al., 2024), and less pride in eating pathology (Brede et al., 2024).

Given the high prevalence of ED in autistic females, the high prevalence of autism or autistic traits in females with ED (C. M. Brown & Stokes, 2020; Westwood & Tchanturia, 2017; Wentz et al., 2005; Zhang et al., 2022), and the overlap between autistic traits and ED characteristics (Nistico et al., 2022; see also Supplementary Materials), some authors have proposed that anorexia could represent a female-specific manifestation of autism (Carpita et al., 2022; Odent, 2010). Further research is needed for clarification, and improved screening for autism is recommended in individuals with anorexia nervosa (Beygui & Cascio, 2022).

Autistic characteristics were also directly or indirectly associated with anxiety and depressive disorders, with sex differences. Autistic female children and adolescents had more anxiety symptoms than males at low levels of autistic characteristics, but did not differ from males on anxiety symptoms at high levels of autistic characteristics (Long et al., 2024). This study also showed that the effect of autistic characteristics on depressive symptoms was similar for autistic males and females. RRBs were more elevated in autistic children and adolescents with anxiety disorder than in those without (Wodka et al., 2022), and in adults with higher anxiety symptoms (Kuzminskaite et al., 2020). However, this was not observed in females aged 6–12 years, suggesting that RRBs may have served as coping mechanisms for anxiety, but that younger females might have relied on different strategies (Wodka et al., 2022). Alternatively, their anxiety may have manifested differently, with fewer overlaps with RRBs (Wodka et al., 2022). Another study found that certain RRBs, including hair and skin pulling, rubbing, or scratching, were more common in autistic females aged 3–18 years than males (Antezana et al., 2019). While these behaviors may reflect underlying MH issues, they could also be driven by autistic characteristics such as sensory differences. Both sensory sensitivity and low sensory registration were associated with depressed mood in autistic female children and adolescents, consistent with the behavioral withdrawal model of depression (Bitsika, Sharpley, & Mills, 2021).

Social communication difficulties were also linked to elevated anxiety in autistic children (Dubin et al., 2015), particularly in females (Wodka et al., 2022), and to the number of MH diagnoses in autistic adults (Dubreucq et al., 2023). In addition, low social well-being and dissatisfaction with social support in autistic adults contributed to depression (Hedley et al., 2018; Richdale et al., 2023), a relationship mediated by loneliness (Hedley et al., 2018). Loneliness itself was associated with both anxiety and depression and was influenced by the quality and quantity of friendships (Hedley et al., 2018; Mazurek, 2014). However, these associations were not moderated by sex.

Autistic characteristics also influenced suicidality (Bentum et al., 2024), often in conjunction with feelings of burdensomeness and social disconnection (Cleary et al., 2023; Pelton et al., 2023). These factors may contribute to the associations between suicidal behavior and being unemployed or not in a romantic partnership (Kolves et al., 2021). Again, these associations were not moderated by sex (Hedley et al., 2018; Pelton et al., 2023).

Finally, one specific characteristic, brooding, which is not inherently autistic but often elevated in autistic people, may further reinforce these associations. It was found to increase depressive symptoms in autistic individuals, particularly females, who, unlike their male counterparts, did not seem to benefit from the protective effects of self-focused attention (Burns et al., 2019).

#### Intellectual and language abilities

ID was more prevalent in autistic females than in autistic males in PopReg studies (15–38% vs. 11–25%; e.g., Angell et al., 2021; Dillon et al., 2023; Hsu et al., 2024; but Rødgaard et al., 2021) and was identified as a potential protective factor against MH issues. Indeed, the absence (vs. presence) of ID and higher IQ (vs. lower IQ) in autistic individuals has been associated with higher likelihood of anxiety disorders (Salazar et al., 2015; Wodka et al., 2022) and higher anxiety symptoms (Dubin et al., 2015; Long et al., 2024; Mayes et al., 2011). One PopReg study suggested that this association was particularly pronounced in female children and adolescents (Long et al., 2024). In contrast, among autistic people with ID, this sex difference was not observed either in children and adolescents (Magiati et al., 2016) or adults (Tsakanikos et al., 2011).

For specific cognitive abilities, studies in autistic children found that anxiety symptoms increased with verbal IQ, although no sex differences were observed (Mayes et al., 2011), while social anxiety symptoms decreased with higher matrix reasoning scores in autistic female children and adolescents (Bitsika & Sharpley, 2024). These findings suggest that anxiety in autism was more closely related to verbal than to non-verbal abilities, although further investigation is warranted. In addition, being verbal was associated with higher scores on the emotional dysregulation dysphoria subscale (reflecting low positive affect and sadness) in autistic children and adolescents (Wieckowski et al., 2020).

Higher cognitive abilities were also related to heightened vulnerability to depressive disorders (De-la-Iglesia & Olivar, 2015; Greenlee et al., 2016) but not symptoms (Long et al., 2024), or at least rendered them more detectable. In contrast to the general and overall autistic populations, where females are more at risk for depression, one study found that autistic males aged 30–49 years were the most vulnerable group among adults with ID (Saez-Suanes, Garcia-Villamisar, & del Pozo-Armentia, 2023).

Besides having fewer anxiety and depressive disorders, autistic individuals with ID exhibited more behavioral and conduct problems (Kerns et al., 2020). In addition, lower IQ, particularly verbal IQ, was associated with increased risks of self-injurious behaviors and severe self-harm, both at home and in clinical settings (Cohen et al., 2010; Handen et al., 2018; Lai, Saunders, et al., 2023). While a study in autistic children and adolescents did not report sex differences (Handen et al., 2018), another in adults found that autistic males were more likely to direct physical aggression toward others, whereas autistic females were more likely to direct it toward themselves (Cohen et al., 2010). Autistic females with ID were also more likely than their male peers to make self-deprecating statements. These findings suggest that MH difficulties may play a specific role in self-destructive behaviors among autistic females with ID (Cohen et al., 2010).

Findings regarding suicidality in autistic individuals with ID remained mixed. Several studies reported reduced rates of suicidality (Holden et al., 2020), suicide attempts (Hirvikoski et al., 2020), and suicide deaths (PopReg study: Lai, Saunders, et al., 2023; Meta-analysis: Santomauro et al., 2024) in those with ID compared to those without. In contrast, other studies suggested that while ID, particularly lower verbal IQ, was associated with less suicidal ideation or talk of self-harm, it was linked to more suicide attempts (Hand, Benevides, et al., 2020; McDonnell et al., 2020; for a scoping review see Cleary et al., 2023). Nonetheless, the risk of suicide attempts in autistic individuals remained elevated compared to their non-autistic peers, regardless of ID status (Hirvikoski et al., 2020). There was also some evidence that risks of suicide attempt may have been higher in autistic females than males, with or without ID, although formal analyses of sex-by-group differences were lacking (Hirvikoski et al., 2020).

There were limited findings for other MH conditions. One study reported higher rates of schizophrenia spectrum disorders and personality disorders in autistic males with ID (Tsakanikos et al., 2011). In a sample of autistic children and adolescents with ID, no sex differences were observed in overall MH symptoms (Gobrial, 2019).

### ADHD

ADHD is another neurodevelopmental condition frequently co-occurring with autism. PopReg studies in adolescents and adults reported a lower prevalence of ADHD in autistic females (2.6–55%) than males (2.4–61%; Angell et al., 2021; Croen et al., 2015; Dillon et al., 2023; Hand, Angell, et al., 2020; Hsu et al., 2024; Rødgaard et al., 2021; Vadukapuram et al., 2022). However, another PopReg study in children (Stacy et al., 2014) and studies including adults indicated no sex differences (Jadav & Bal, 2022; Kerns et al., 2020). Similar outcomes emerged from ClinCom studies, reporting a higher risk in autistic males during childhood (Salazar et al., 2015) but no sex differences in adulthood (Lugnegard et al., 2011). The discrepancy between childhood and adulthood findings potentially reflects the under-recognition of ADHD in autistic female children.

Autistic individuals with ADHD were at increased risks for MH issues (Accardo et al., 2024; Hirvikoski et al., 2020; Zhao et al., 2024). Specifically, autistic female children and adolescents with ADHD were more vulnerable to anxiety problems, followed by autistic males with ADHD, although sex differences were not always statistically significant (Accardo et al., 2024; Zhao et al., 2024). Depression was also more prevalent among autistic children and adolescents with ADHD than those without, and was either higher in females (Zhao et al., 2024) or similar across sexes (Accardo et al., 2024). Nevertheless, the latter showed an increased risk of depressive disorders in autistic males with ADHD, which was not observed in females. Overall, ADHD might heighten the risks of anxiety and depressive disorders in autistic children and adolescents, while also reducing sex differences in the likelihood of being diagnosed with these disorders. Finally, a PopReg study showed that ADHD was associated with increased risks of suicidal behavior in autistic people, with autistic females particularly at risk (Hirvikoski et al., 2020).

#### Sleep

Sleep-wake disorders were elevated in autistic compared to non-autistic people (Estes et al., 2023; Hand, Angell, et al., 2020; Lai et al., 2019; May, Cornish, Conduit, et al., 2015), especially among autistic females compared to non-autistic females (Henderson et al., 2023). Sleep diagnoses or problems increased with age in autistic people (Jadav & Bal, 2022). PopReg studies reported significantly higher rates in autistic females (2–40%) than in males (1.2–38%; Angell et al., 2021; Dillon et al., 2023; Hand, Angell, et al., 2020; Jadav & Bal, 2022; Martini et al., 2022; Vadukapuram et al., 2022), though one study found no sex differences (Rødgaard et al., 2021). ClinCom studies also reported more sleep difficulties in autistic female children (Estes et al., 2023; Hartley & Sikora, 2009) and young adults (Williams & Gotham, 2022) than males, including sleep anxiety, bedtime resistance, shorter sleep duration, and sleepiness, which could all be interrelated (Estes et al., 2023). In addition, autistic adult females had overall poorer sleep quality (Charlton et al., 2023; Migliarese et al., 2020) but similar sleep efficiency (Henderson et al., 2023) than autistic males. This sex-related pattern was observed in non-autistic people as well (Charlton et al., 2023; but see Henderson et al., 2023).

Sleep-wake disorders may result from, while also exacerbating, MH issues, with a stronger link between objective sleep efficiency and the lifetime history of MH illness or distress history in autistic than in non-autistic adults (Henderson et al., 2023). However, findings were mixed about the associations between sleep and MH in autistic people. While one study found no link between anxiety and sleep quality in autistic adults (Migliarese et al., 2020), two others reported significant associations in both adults (Charlton et al., 2023) and children (May, Cornish, Conduit, et al., 2015). Anxiety and perceived stress were linked to sleep disturbance and daytime dysfunction in adults (Charlton et al., 2023), and sleep disturbance predicted anxiety one year later in children (May, Cornish, Conduit, et al., 2015). In another study, a positive association between anxiety symptoms and sleep difficulties was observed only in autistic male children (Estes et al., 2023).

While sleep quality and fatigue were central contributors to depression in autistic adolescents and young adults, more so than in their non-autistic peers (Montazeri et al., 2020; Richdale et al., 2023), sleep problems were not more prevalent among ever-depressed autistic children and adolescents (Greenlee et al., 2016), with no sex differences reported in either study. In contrast, depression predicted poorer sleep quality in autistic adults, especially among females, who showed notably reduced sleep duration (Migliarese et al., 2020). This sex effect may reflect an innate vulnerability or stem from the higher prevalence of internalizing disorders in autistic adult females. The authors proposed that sleep problems could constitute a sex-specific autism phenotype, either as a direct manifestation or secondary to the high depression rates observed in females.

#### Physical health

PopReg studies reported a higher incidence of physical health issues in autistic compared to non-autistic individuals (Hand, Angell, et al., 2020), and in females compared to males (Croen et al., 2015; DaWalt et al., 2021). Autistic females appeared particularly at risk for epilepsy, metabolic and autoimmune disorders, thyroid disease, diabetes, and gastrointestinal problems (Angell et al., 2021; Croen et al., 2015). While no sex differences in somatic symptoms existed in childhood (Jolly et al., 2020), autistic females reported such symptoms more often than autistic males in adulthood (54% vs. 19%; Williams & Gotham, 2022). Chronic pain, whether localized or widespread, was also especially prevalent among autistic females (Asztély et al., 2019; Williams & Gotham, 2022). These physical health issues in autistic females potentially contribute to their lower physical health-related quality of life compared to non-autistic females, a disparity not seen among males (Braden et al., 2021).

These physical health difficulties may also contribute to the MH vulnerability of autistic females, as physical and mental health are often interrelated. For instance, somatic symptoms were associated with greater depression, anxiety, autistic traits, and reduced quality of life in autistic adults (Williams & Gotham, 2022). Similarly, autistic children and adolescents with more seizures and gastrointestinal problems were more likely to experience depression (Greenlee et al., 2016). Gastrointestinal issues and ear infections were linked to increased sleep problems and self-injurious behaviors in autistic adults with ID (Cohen & Tsiouris, 2020). In particular, for autistic females, physical discomfort was a trigger for self-injury and aggression, possibly due to difficulties recognizing and/or expressing discomfort, leading to self-injury as a potential coping mechanism (Cohen & Tsiouris, 2020). However, despite reporting poorer overall health, autistic females with chronic pain did not report poorer MH than those without chronic pain (Asztély et al., 2019). While pain ratings in autistic children were linked to suicidal ideation (McDonnell et al., 2020), no significant association was found between physical comorbidities and suicide attempts in autistic adolescents or adults (Kolves et al., 2021).

#### Filling the gap: towards a mechanistic understanding of sex differences

Despite growing evidence of sex differences in the prevalence of MH difficulties and modulating factors, their underlying mechanisms remain insufficiently understood. Below we highlight emerging directions uncovering the mechanisms.

#### Biological contributions

Genetic factors may constitute a primary source of sex-based differences. Autistic females exhibit a higher genetic load associated with autism than males. This may increase their vulnerability to MH conditions, as many autism-related genetic variants are associated with heightened risks for psychiatric disorders (DaWalt et al., 2021; Niarchou et al., 2023; Rødgaard et al., 2021; Wright et al., 2023). This hypothesis aligns with findings showing that autism polygenic scores are not only linked to autism diagnosis but also to mood disorders and depression in autistic females, an association not observed in autistic males (Niarchou et al., 2023). However, differences between sexes in these associations were not statistically significant after accounting for the baseline prevalence of each condition, suggesting a broader female-specific vulnerability.

Sex-related biology can shape neural functioning, including MH susceptibility, in various ways. For example, sex hormones influence the neuroendocrine system, which in turn regulates emotional and stress responses. One particularly relevant pathway is the hypothalamic–pituitary–adrenal (HPA) axis, which plays a key role in stress responses. Autistic children and adolescents showed higher evening cortisol levels than non-autistic individuals, a pattern associated with poorer response to change, and potentially reflecting chronic hyperarousal and cumulative stress, contributing to a hyperactive HPA axis (Corbett, McGonigle, et al., 2024). Notably, both autistic and non-autistic females exhibited higher evening cortisol than males, although no association was found between evening cortisol and depressive symptoms in females, a link not examined in males (Corbett, McGonigle, et al., 2024). Other studies, however, reported associations between depressive symptoms in autistic female children and adolescents and dysregulated cortisol awakening response, in the opposite direction to what is typically observed (Sharpley et al., 2016), as well as with morning cortisol levels (Sharpley et al., 2021). The authors contrasted these findings with their earlier study in autistic male children and adolescents, where anxiety, rather than depression, was associated with cortisol changes. In males, these changes involved diurnal fluctuations in cortisol concentration, rather than cortisol awakening response (Sharpley et al., 2016). Thus, chronic stress might affect the HPA axis and MH differently in autistic females and males, possibly contributing to depression in autistic females and anxiety in autistic males.

Sex hormones may also have a direct influence on neural functioning. Due to heightened sensitivity to hormonal changes, autistic females may be particularly vulnerable to sex hormone fluctuations. This has been illustrated in studies reporting significant challenges experienced by autistic female children and adolescents during the premenstrual and menstrual periods (Groenman et al., 2022; Lee, 2004; Lundin et al., 2021; Obaydi & Puri, 2008; Skinner et al., 2005; Vijapura et al., 2014; see section Sex-related physiology). Sensitivity to hormonal shifts was also described by an autistic adult female in a qualitative study on pregnancy (Hampton et al., 2023). In addition to increased sensitivity, or as an alternative explanation, the physiological hormonal balance in autistic females may differ from that of non-autistic females (Groenman et al., 2022). The authors speculated that highly irregular fluctuations in estrogen could account for the more frequent and prolonged menopausal challenges observed in autistic females.

#### Social contributions

Social expectations, especially gendered norms that pressure females to display greater social competence and affiliation, may further burden autistic females, and may interact with hormonal changes to heighten MH difficulties, especially during adolescence. Social engagement typically increases during this period, particularly in females, who are more likely than autistic males to engage in social interactions in late adolescence and adulthood (Chen et al., 2017). However, autistic females’ struggle with social integration may contribute to their greater risks, compared to autistic males, of being ignored or experiencing victimization and bullying during childhood and adolescence (31% vs. 23%; R. E. Adams et al., 2020; Holden et al., 2020). Such social exclusion and bullying were associated with negative MH outcomes, including emotional distress, depression and anxiety, particularly in autistic adolescent females compared to males (Greenlee et al., 2020), sometimes manifesting as compulsive behaviors (Fowler & O’Connor, 2021) and increased suicidality (Holden et al., 2020).

To avoid exclusion and its detrimental effects, autistic individuals often engage in camouflaging, noted at higher levels in females (Cassidy et al., 2018; Cook et al., 2021; Lai et al., 2017), although the strategies were not qualitatively different from males (L. Bradley et al., 2021). However, the sustained efforts to mask autistic traits can lead to MH challenges (Cook et al., 2021; Lai, Amestoy, et al., 2023). Camouflaging was described as exhausting and isolating by autistic adolescents and adults, with harmful effects on physical and MH (L. Bradley et al., 2021; Halsall et al., 2021). Higher camouflaging was associated with increased symptoms of anxiety, depression, and stress in adolescents regardless of being autistic or not, with stress symptoms specifically linked to female sex (Bernardin et al., 2021). In autistic adolescent females, uncertainty about the success of camouflaging may also trigger anger and anxiety according to qualitative interviews (Halsall et al., 2021). In autistic adults, camouflaging correlated with anxiety symptoms (Hull et al., 2021), suicidality (Cassidy et al., 2018), and ED symptoms (S. Bradley et al., 2024). While self-reported camouflaging in autistic adults correlated with depressive symptoms (Hull et al., 2021), this link appeared in males only when using a discrepancy method partly based on external observation (Lai et al., 2017). One study showed that the link between camouflaging and depressive symptoms in autistic people was mediated by emotion regulation difficulties and perceived stress, with stronger effects in females (McQuaid et al., 2024).

Behavioural manifestations of challenges related to camouflaging and MH difficulties in autistic female children and adolescents are often more apparent at home (Chandler et al., 2016; Halsall et al., 2021), as they try to hide their difficulties at school (Halsall et al., 2021). This is reflected in anxiety and depression scores, which more often exceed clinical cut-offs when rated by parents than by teachers, particularly for females (Chandler et al., 2016). Teacher-reported generalized anxiety symptoms were higher in autistic males than females (D. Adams et al., 2018), supporting the idea that females may also camouflage MH symptoms. Effortful control used by females to camouflage autistic traits and MH struggles may itself negatively affect MH (Halsall et al., 2021). Finally, some studies reported higher parent-rated than self-rated anxiety (Bitsika, Sharpley, Mandy, et al., 2021; Bitsika & Sharpley, 2020), which could reflect either parental overestimation or a reduced symptom awareness in autistic females, in whom anxiety may be experienced as an intrinsic part of the self (Bitsika & Sharpley, 2020).

### Better support: Tailoring care

Being aware of MH issues in autistic females, as well as the contributing factors and underlying mechanisms, is essential for identifying downstream consequences and developing tailored and effective MH support.

#### Downstream consequences

MH difficulties, which are more prevalent in autistic females, often act as a gateway to additional challenges in autistic individuals and can trigger a cascade of adverse outcomes. First, MH issues may exacerbate autism-related difficulties, potentially in sex-specific ways. In a study of autistic and non-autistic adolescent females, higher internalizing and externalizing symptoms were linked to lower social competence, although the association with internalizing symptoms was significant in non-autistic females only (Jamison & Schuttler, 2015). Among autistic females, the correlation between internalizing symptoms and social competence was small and non-significant, while the correlation with externalizing symptoms was moderate and significant. This suggests that, among autistic adolescent females, internalizing symptoms might be less related to social difficulties than externalizing symptoms. Supporting this, one study found that having symptoms in “sex-incongruent” domains (e.g., internalizing in males, aggression in females) moderated the association between emotional dysregulation and social success in autistic children and adolescents (Neuhaus et al., 2019). When emotional dysregulation manifested as irritability and aggression, it blunted the facilitative effects of social motivation on social skills, for females more so than males, thus impairing social participation and well-being. At the extreme, a study showed that externalizing behaviors and co-occurring diagnoses such as ADHD or psychotic disorders significantly doubled the contact with the criminal justice system prior to autism diagnosis (Blackmore et al., 2022). Nevertheless, contact with the justice system remained lower among autistic adult females compared to males (11% vs. 31%).

Another downstream consequence of MH difficulties was the high likelihood for autistic individuals to receive one or more psychotropic medications (Barnette et al., 2019; Caplan et al., 2022; Jobski et al., 2017). This occurred more frequently than in non-autistic people (E. Bradley & Bolton, 2006), especially among those with co-occurring ADHD and/or anxiety (Wodka et al., 2022), ID, aggressive behavior, or seizures, with no sex differences reported (Barnette et al., 2019; Caplan et al., 2022). On the contrary, a PopReg study found that a higher proportion of autistic male children and adolescents took psychotropic medication compared to autistic females (43% vs. 39 %; Wodka et al., 2022). These inconsistencies may reflect prescription differences: autistic males were more often prescribed stimulants, antipsychotics, and polypharmacy, whereas autistic females were more often prescribed antidepressants, anxiolytics, hypnotics-sedatives, and antiepileptic medications (Caplan et al., 2022; Jobski et al., 2017; Tsakanikos et al., 2011). Further, a PopReg study (DaWalt et al., 2021) indicated that autistic adult females show greater healthcare utilization than males or non-autistic peers for psychiatric, nutritional, neurological, and sleep-related conditions, likely reflecting a greater burden of co-occurring conditions, diagnostic complexities, and/or differences in treatment response. These sex-specific considerations are particularly important given the limited data on differential medication efficacy and side effects in autistic people, as highlighted in a narrative review with several examples (Green et al., 2019). Antipsychotics may pose unique challenges for autistic females, such as the potential side effects of hyperprolactinemia that are more frequent and particularly distressing in females (e.g., menstrual irregularities and galactorrhea). Valproic acid increases the risk of polycystic ovary syndrome, to which autistic females may already be more vulnerable. In addition, certain psychotropic medications may reduce the effectiveness of hormonal contraception, another concern often overlooked.

Furthermore, in an online study, autistic adolescents and adults were three times more likely than non-autistic individuals to use recreational psychotropic substances to cope with MH symptoms, while non-autistic individuals were more likely to use them for social purposes (Weir et al., 2021). Autistic individuals also reported using these substances as self-medication for “physical” symptoms such as sleep or eating difficulties; however, the frequency of this use did not differ significantly between autistic and non-autistic individuals (Weir et al., 2021). Notably, receiving an autism diagnosis was reported to be relevant for some individuals in reducing or stopping their substance use. Although sex differences were not directly examined, the study found that, unlike autistic males, who were less likely to use substances than their non-autistic counterparts, autistic females showed similar rates of use to non-autistic females, suggesting that they may be less protected from engaging in substance use than autistic males. MH difficulties significantly impair quality of life, both directly and indirectly (e.g., through medication side effects, increased social difficulties). In autistic children, elevated anxiety was linked to lower child (and parent) quality of life, regardless of sex (D. Adams et al., 2020). Among autistic adolescent females, higher internalizing and externalizing symptoms were associated with lower quality of life (Jamison & Schuttler, 2015). Autistic adults’ health-related quality of life was lower than that of non-autistic adults (Braden et al., 2021). Yet, in autistic adult females, older age was associated with improved MH-related quality of life (Braden et al., 2021). Other studies showed that autistic males reported lower social quality of life than females, while autistic females reported lower physical, psychological, and environmental quality of life than males (Lawson et al., 2020). This aligns with findings suggesting that both being female and having MH difficulties were independent and additive predictors of poorer quality of life in autistic adults (Mason et al., 2018).

Finally, MH issues may increase the emergence of other MH issues. For example, autistic adult females with ED reported more co-occurring MH issues than non-autistic peers, particularly social anxiety (Brede et al., 2024). Self-reported social anxiety in autistic female children and adolescents was also linked to their total scores on a questionnaire targeting ED in autistic people (Sharpley & Bitsika, 2022). In addition, while one ClinCom study in autistic children did not find a significant association between internalizing or externalizing symptoms and suicidal ideation (Hunsche et al., 2020), many other studies consistently reported that psychiatric conditions, such as mood and anxiety disorders, ED, PTSD, and emotional dysregulation, were significant correlates of suicidality in both autistic males and females (Arwert & Sizoo, 2020; Bentum et al., 2024; Cleary et al., 2023, p. 202; Conner et al., 2020; Costa et al., 2020; Hand, Benevides, et al., 2020; Holden et al., 2020; Kolves et al., 2021; Lai, Saunders, et al., 2023; McDonnell et al., 2020; Storch et al., 2013; Zhang et al., 2022). Cassidy et al. (2018) also highlighted unmet support needs as a risk factor, although its impact was diminished when psychiatric diagnoses were controlled for (Hirvikoski et al., 2020; Lai, Saunders, et al., 2023). These findings underscore the importance of promptly identifying and addressing MH concerns, not only to improve immediate well-being but also to prevent a broader set of downstream negative consequences. Yet, suicide prevention initiatives and training remain underdeveloped in this population (Cleary et al., 2023).

#### Support

Lai, Amestoy, et al.’s (2023) viewpoint highlights the health inequities experienced by autistic females, spanning social, institutional, research, and policy domains. An Australian study found that 73% of autistic females and 65% of autistic males (a non-significant difference) reported needing professional support for MH and well-being, though this need was unmet in approximately 30% of cases (Baldwin & Costley, 2016). Financial limitations and a lack of affordable or funded services were identified as key barriers by autistic adult females in a qualitative study (Tint & Weiss, 2018). Moreover, some professionals appear unable or unwilling to adapt their interventions to the needs of autistic females, limiting their effectiveness (Tint & Weiss, 2018).

Kelly et al. (2024) offer recommendations for improving care for neurodivergent females in a narrative review, emphasizing active listening, allowing more time, ensuring consistency and predictability, adapting communication (e.g., offering both oral and written information), and enhancing interprofessional collaboration. While these practices may benefit all autistic individuals, tailored approaches are also needed to address MH vulnerabilities during critical life stages of autistic females, such as premenarchal counselling and hormonal management during puberty to alleviate distress (Fei et al., 2021). Likewise, nurses and midwives with autism-specific training can reduce anxiety by clearly communicating what is happening before and during childbirth (Donovan, 2020).

Autism-informed approaches are also critical. Treatment for depression should consider the role of sensory profiles in the development and experience of depression in autistic females (Bitsika, Sharpley, & Mills, 2021). Similarly, it is essential to understand how autistic traits, such as sensory sensitivities, rigidity, and a strong preference for routine and predictability, contribute to the onset, persistence, and severity of disordered eating behaviors (Beygui & Cascio, 2022; Schroder et al., 2023). Treatment for ED is often hindered by a lack of autism understanding (Babb et al., 2021; C. M. Brown & Stokes, 2020; Schroder et al., 2022, 2023). Although autistic females access more care than non-autistic females, they report less benefit (Babb et al., 2022; Schroder et al., 2023). For example, dietetic interventions may be less effective when sensory sensitivities are not adequately understood, and Cognitive Behavioral Therapy (CBT) and cognitive remediation may show limited efficacy due to reduced emphasis on weight and body shape concerns in autism-related ED (Babb et al., 2021, 2022; Schroder et al., 2023). Conversely, therapies focused on emotion regulation, such as Dialectical Behavior Therapy (DBT), were perceived as more beneficial (Babb et al., 2021). However, these findings mainly come from qualitative studies, and no research to date has evaluated the efficacy of these or other adapted therapies for individuals with co-occurring autism and ED. Therefore, adapting treatments remains a key priority to improve interventions and staff training for better outcomes in this population (Beygui & Cascio, 2022; Lai, 2023; Zhang et al., 2022).

Although CBT may be less effective for ED, it can nevertheless reduce psychological distress in some autistic adults, with one study finding 37% showing reliable improvement, independent of sex or age at autism diagnosis (Blainey et al., 2017). However, those diagnosed in adulthood presented greater psychological distress and were more likely to require longer treatment, which may be particularly relevant to autistic females, who tend to be diagnosed later in life.

Given the heightened emotional dysregulation in autistic females and its link with anxiety (Saez-Suanes, Garcia-Villamisar, & Pozo Armentia, 2023; Weiner et al., 2023), interventions targeting emotion regulation may be particularly suitable. A study of autistic adults eligible for DBT pointed to sex-specific factors underlying emotional dysregulation in autism, underscoring the importance of targeting specific aspects in treatment, such as addressing alexithymia specifically in autistic females (Weiner et al., 2023).

Considering that autistic females experienced higher levels of activity-related and event-related stress, interventions targeting stress management may help mitigate the negative impacts and reduce MH symptoms (Ilen et al., 2024). A heightened affective response to daily stressors was also linked to the use of fewer adaptive coping strategies (e.g., acceptance, refocusing, putting things into perspective) and more non-adaptive ones (e.g., rumination, self-blame, catastrophizing), which may contribute to the severity of MH difficulties (Ilen et al., 2024). Accordingly, mindfulness-based stress reduction and psychoeducation interventions might be beneficial for autistic individuals, with initial evidence suggesting a greater improvement in MH-related quality of life among autistic adult females compared to males (Braden et al., 2021).

Autistic females also reported lower self-compassion than autistic males or non-autistic individuals, which was associated with greater anxiety and depression (Galvin & Richards, 2023). Cultivating self-compassion may therefore be another promising avenue for improving the MH of autistic females.

Other approaches have been explored in autistic females; however, available data are often limited to case-studies. For instance, the Girls Night Out program specifically targets the socio-emotional health of autistic adolescent females, with sessions held in community settings and involving non-autistic peers as mediators. The intervention consists of 2-hour weekly sessions over 12 to 16 weeks. Preliminary findings from 34 autistic adolescent females showed significant improvements in self-reported internalizing symptoms and quality of life (Jamison & Schuttler, 2017). One case study also highlighted the benefits of the Piece it Together wellness program (twelve 90-minute classes focused on exercise, nutrition, socialization, and stress reduction) in reducing anxiety and enhancing social engagement in an autistic young adult female (Spratt et al., 2019). These results align with findings linking adherence to lifestyle guidelines (physical activity, sleep, screen time) with lower odds of anxiety and depression in autistic children with ADHD (Zhao et al., 2024). Externalizing Metaphors Therapy, which encourages clients to externalize anxiety through metaphor, helped reduce symptoms to be more manageable in a 15-year-old autistic female within four sessions (McGuinty et al., 2018). A case study using a humanoid robot reported reduced social anxiety in a 22-year-old autistic female with social anxiety disorder and an interest in technology, who had not responded to extensive CBT; she was then able to greet her teacher and classmates for the first time (Yoshida et al., 2022). Lastly, deep brain stimulation of the nucleus accumbens in a minimally verbal autistic adult female with severe OCD led to improvements in both OCD symptoms and unexpected gains in communication and self-advocacy (Doshi et al., 2019); this invasive approach raises ethical concerns, and generalizability, safety, and accessibility remain to be assessed. Research into innovative interventions for autistic females with high support needs, often underrepresented in studies, is urgently needed.

## Discussion

The objective of this rapid review was to map sex differences in the prevalence and presentation of MH conditions in autistic people, explore influencing factors and underlying mechanisms, and identify tailored support strategies. A total of 218 studies were included, combining PopReg and ClinCom studies, together with literature reviews, meta-analyses, and relevant case studies, commentaries and viewpoints, to offer a comprehensive synthesis.

Regarding the *objective 1* of mapping sex differences in the prevalence and presentation of MH conditions in autistic people, our findings highlight a large body of literature. Across studies, autistic females were at greater risk than autistic males for experiencing at least one MH disorder, a pattern also observed in the non-autistic population. Autistic females seem particularly vulnerable to internalizing conditions compared to males, as confirmed by our exploratory meta-analytic syntheses on PopReg studies. Indeed, the quantitative syntheses showed a higher likelihood of anxiety disorders, mood disorders in general and more specifically depressive and bipolar disorders, ED, and OCD in autistic females than males. This pattern was also observed for schizophrenia spectrum and other psychotic disorders, personality disorders, and suicide attempts (but not suicide deaths), while findings were more mixed for SUD.

Overall, the findings imply a generally additive vulnerability: autistic individuals already face higher MH vulnerability than non-autistic individuals, and this risk may further increase in females. However, some studies suggest that the odds ratio of having a MH condition in autistic vs. non-autistic individuals is usually higher in males than in females, except for schizophrenia and other psychotic disorders. This potentially reflects sex differences in the baseline prevalence observed in the general population, which is higher in males for schizophrenia (Aleman et al., 2003), contrary to internalizing disorders that are more prevalent in females.

Inconsistencies exist in prevalence estimates across studies. This is likely explained by differences in sample size and ascertainment methods. PopReg studies reported more consistent sex differences than ClinCom studies. In addition, ICD-code-based diagnoses in population-based registries sometimes yielded lower rates than self- or caregiver-reported diagnoses, especially for OCD. The opposite trend was found for schizophrenia spectrum and other psychotic disorders in autistic adults. These discrepancies may arise from several reasons: clinicians might mention symptoms to their clients without formally registering a diagnosis, while self-reports might reflect a broader interpretation of diagnostic labels by the individual or a tendency to report less stigmatized diagnoses (Hazell et al., 2022). Higher self-reported diagnoses than population-based prevalence rates are also sometimes due to subthreshold symptoms or the presence of other confounding conditions (Sordo Vieira et al., 2022). Some differences might also arise from selection bias, particularly affecting online surveys, which can lead to distortion in effect estimates (Rubenstein & Furnier, 2021). Finally, secular trends and cohort effects could also play a role. For instance, the two PopReg studies that did not report sex differences in anxiety were also the oldest (Ahmedani & Hock, 2012; Stacy et al., 2014). This may reflect a changing diagnostic and overall autistic population profile over time, with more autistic females being identified today who may have previously been missed.

It is also important to note that studies exploring sex differences at the symptom level, particularly for anxiety and depression, have reported more mixed findings than those examining clinical diagnoses. Specifically, they showed either no sex differences or increased symptom severity in females. While clinical diagnoses reflect manifestations that meet thresholds of functional impairment, symptom-based assessments may be more sensitive to subclinical distress. Mixed findings across symptom-level studies may also stem from methodological variability and the limited reliability and validity of commonly used assessment tools in autistic people (Halvorsen, et al., 2025; Kim & Lecavalier, 2022). These observations highlight how methodological differences in MH ascertainment may substantially influence the reported patterns of sex differences. In addition, they might contribute to misdiagnosis, delayed diagnoses and inadequate care (Au-Yeung et al., 2019).

### Future directions

While our work represents a first step toward a systematic synthesis of sex differences in MH in autistic people, with the pressing need of more empirical studies that incorporate the investigation of sex differences, future work should conduct systematic reviews and meta-analyses that clearly separate diagnostic categories and symptom-level analyses. Current evidence on MH symptomatology remains mostly descriptive and heterogeneous, making it difficult to draw firm conclusions. In addition, to improve clarity and utility of findings, future empirical studies should aim for harmonized reporting of sex-disaggregated rates. The current review also highlighted underexplored areas, such as geriatric MH conditions or Post-Traumatic Stress Disorder (see Supplementary Materials). From a clinical perspective, our review invites specific attention toward the MH presentation of autistic females, whose symptoms might sometimes be masked or mistaken. Importantly, future work should address barriers to MH support faced by autistic individuals by refining and validating current tools through participatory approaches involving autistic people (Alexandrovsky et al., 2025; Benevides et al., 2020), to improve their accuracy and clinical utility (Cook et al., 2024; O’Connor et al., 2024).

For *objective 2*, we reviewed modulating factors that may influence sex differences in the MH of autistic people. These include sex-related physiology, gender-related experiences, age, age at autism diagnosis, autistic characteristics, and co-occurring conditions such as ID, ADHD, sleep problems, and physical health issues. These factors likely contribute to a cumulative and modified risk for MH issues, especially in autistic females.

Female-specific physiology, such as menstruation, pregnancy, and menopause, can act as MH stressors, for example due to associated physical changes and pain, although little research has been conducted. This mirrors the lack of research on female health in general, particularly on menopause (Dong et al., 2023; Hallam et al., 2022) and menstruation, whose psychological aspects have been considered in research only recently (Gouvernet & Brisson, 2024). This gap further highlights the broader lack of attention to autistic individuals as potential parents, limiting our understanding of how pregnancy and parenting influence MH outcomes. Diagnostic delays in autistic females, often occurring only after a child’s diagnosis (Pohl et al., 2020), further hinder prospective investigations.

Gender diversity was also associated with elevated MH difficulties. It is more prevalent among autistic than non-autistic populations and might be even more so in AFAB individuals (Bonazzi et al., 2025). However, past research rarely examined the intersections of sex and gender in autism (Strang et al., 2020), and their respective and combined impact on MH. Among autistic AFAB individuals, the rapid and disorienting bodily changes also affect sensory experiences and may prompt gender distress. Understanding how these experiences are related to and different from a stable sense of gender incongruence and the developing personal identity requires careful and empathic clarification. A nuanced exploration is needed to better understand pubertal distress in autistic AFAB individuals and its association with gender dysphoria, to better target MH support and gender-related care (Bo et al., 2024).

ID was associated with a seemingly lower risk of MH conditions such as anxiety, contrary to other factors. However, it was associated with more behavioral or aggressive symptoms, which may be more often self-directed in autistic females. This negative association between ID and MH issues may be related to reduced introspective or verbal capacities, and limitations in how MH is assessed in autistic people with ID and/or language impairments (Peña-Salazar et al., 2022), who may express distress through aggressive behaviors. In particular, self-injurious behaviors in autistic females with ID may be interpreted by others solely through the lens of stereotyped behaviors, without considering them as potential indicators of emotional distress or physical illnesses, which are in fact elevated in autistic females (Kassee et al., 2020).

### Future directions

Modulating factors of sex differences in MH among autistic people remain under-researched and rarely assessed specifically. Future research should identify these factors, characterize their complex relationship with autism and co-occurring MH conditions, and clarify the directionality of their influences (van der Putten et al., 2025). Understanding these mechanisms and their interactions with sex would help identify actionable levers for clinical support. Large-scale longitudinal designs are needed to capture the evolution of MH conditions across the lifespan, considering developmental milestones especially those key to female health (e.g., puberty, maternity, menopause). These studies should also integrate other modulating variables such as co-occurring conditions (e.g., ADHD, ID, physical health issues, sleep problems) and gender-related experiences to capture the complex intersections affecting MH. Advanced multivariate modeling approaches could help clarify how sex, gender, and other modulators interact to shape MH trajectories in autistic people. Finally, substantial work is needed to adapt MH assessments for individuals with ID (Kim & Lecavalier, 2022), in order to better detect and address distress, particularly in females.

*Objective 3* was to explore potential mechanisms. This review underscores the paucity of mechanistic research that could clarify sex differences in the MH of autistic people. Biologically, genetic loads and hormonal variations associated with the female sex, as well as atypical hormonal profiles in autistic females (Gasser et al., 2020; Ingudomnukul et al., 2007; Pohl et al., 2014), may play key roles in the MH vulnerability of autistic females, particularly around reproductive transitions. In particular, atypical estrogen reactivity and sex-differentiated dysregulation of the HPA axis might contribute to increased stress reactivity and MH vulnerability, especially in autistic females (Bitsika, Sharpley, Mandy, et al., 2021; Groenman et al., 2022).

These biological mechanisms likely interact with social experiences. Females often face higher social demands than males due to gendered social expectations imposed on them (Leaper and Friedman, 2007), and many respond with sustained camouflaging efforts. Camouflaging has both cognitive and emotional cost to autistic people, and is associated with exhaustion, anxiety, and depression (Zhuang et al., 2023). As such, social expectations and camouflaging may, hypothetically, represent a pathway through which gendered expectations produce disproportionate MH burdens for autistic females, although the links between camouflaging and MH remain to be clarified (Khudiakova et al., 2025).

### Future directions

Research should address how biological and social mechanisms influence MH outcomes in autistic people, especially females. For instance, there is a need to understand the impact of hormonal variations and treatment on MH in autistic females, with studies that are sufficiently powered and include both hormonal and MH measures. Studies on the impact of contraception and hormonal replacement therapy on quality of life and MH are required (Rynkiewicz et al., 2025). It is also critical to design studies to clarify the interactions of biological and social-experiential effects on the MH of autistic females, for which mixed-method research could be particularly helpful.

Our *objective 4* was to synthesize the impact of unmet MH needs and the options for tailored MH care for autistic females. The urgency is clear as the downstream consequences of MH needs in autistic people are substantial and include suicidality, excessive psychotropic medications and substance use, and reduced quality of life. Some other downstream effects have not been investigated, such as the relationships between MH and burnout/chronic fatigue, which also appear to be higher in autistic females (Kentrou et al., 2021) and associated with camouflaging (Benatov et al., 2025).

Few studies have addressed tailored support strategies. Early and accurate autism diagnosis in females could be a key intervention point to reduce MH burden (Blainey et al., 2017). This calls for better recognition of the nuanced autism phenotypes associated with sex and gender (Lai et al., 2022). Support strategies vary according to MH concerns and could include CBT, DBT, mindfulness, self-compassion approaches, wellness programs, and psychoeducation around female health. Interventions should also address psychosocial dimensions to enhance social well-being, such as social identity, self-acceptance, and self-esteem. These strategies have rarely been evaluated specifically in autistic females to date.

### Future directions

Research needs to generate more evidence for the effectiveness of the MH support strategies used and their potential variation according to sex. Critically, support should include aspects of female health and be tailored to the specific needs of the autistic individual (Lai, 2023). This could be achieved by more participatory approaches and co-production with autistic people to shape support strategies. Programs and resources that enhance connection and socializing, that are sex-informed and gender-informed, may be especially promising (e.g., https://felicity-house.org/). MH support for autistic people, and specifically for autistic females, is an area requiring more research with rigorous designs.

## Limitations

This rapid review has several limitations. First, not all papers were double screened. Although a group-based calibration process was implemented to align decisions, single-reviewer screening may have introduced inconsistencies. In addition, no formal quality assessment was conducted. These decisions reflect both time constraints and the huge breadth of the review, leading to the choice of a rapid review methodology with exploratory quantitative syntheses, rather than a systematic review and meta-analysis. Second, while focusing on diagnosed autistic females strengthens the reliability of findings, it may exclude undiagnosed individuals, limiting the comprehensiveness and generalizability of findings. Third, certain potentially important moderators (e.g., socioeconomic status, cultural context; see Chen et al., 2017) were not explored due to the absence of relevant findings retrieved during the review, although they could play important roles in shaping MH. Fourth, the 20-year span of included studies may have influenced the characteristics of enrolled participants. Older studies likely included autistic females with higher support needs, whereas newer studies probably captured greater phenotypic diversity. This should be considered when interpreting the narrative synthesis.

## Conclusion

Autistic females face a disproportionate MH burden, with higher rates of internalizing disorders compared to autistic males, most clearly observed in PopReg studies. This rapid review highlights a pattern of additive vulnerability, whereby being autistic and female combine to elevate MH risks. Modulating factors, ranging from sex-related physiology to autism-diagnostic delays, interact across the lifespan but are rarely systematically accounted for in research or clinical practice. Empirical research addressing mechanistic insights and tailored interventions barely exists.

There are urgent needs for sex-informed MH understanding and support strategies, incorporating autistic females’ unique life trajectories including menstruation, pregnancy, and menopause. Research needs to shift toward longitudinal and multi-methods designs that integrate biological and social mechanisms with lived experiences. Participatory approaches are particularly relevant for targeting the MH needs of autistic females and co-developing support strategies. These strategies need to be rigorously evaluated to develop evidence-based MH interventions adapted to autistic people, especially for females. There is also a pressing need to improve MH understanding and assessment in underrepresented groups such as autistic females with ID. Addressing these gaps is not only a research priority but clinical imperatives.

## Data Availability

All data produced in the present study are available upon reasonable request to the authors

## Declaration of generative AI and AI-assisted technologies in the writing process

During the preparation of this work the first author used ChatGPT to perform grammatical checks and to improve readability. After using this tool/service, all authors reviewed and edited the content extensively. We take full responsibility for the content of the published article.

## Acknowledgements

The authors would like to thank Miriam Martini, Lisa Croen, Emily Dillon, and Ericka Wodka for kindly providing additional details about their data upon request.

## Author contributions: CRediT

**Adeline Lacroix:** Conceptualization, Methodology, Data curation, Investigation, Formal Analysis, Writing – original draft, Visualization, Writing – review & editing. **Chih-Chen Tzang:** Data curation, Investigation. **Jennifer Xiofan Yu:** Data curation, Investigation, Writing – review & editing. **Benjamin Koshy-Jacob:** Data curation, Investigation, Writing – review & editing. **Mishel Alexandrovsky:** Methodology, Data curation, Investigation, Writing – review & editing. **Anna Winge-Breen**: Data curation, Investigation. **Terri Rodak**: Methodology, Writing – original draft, Writing – review & editing. **Meng-Chuan Lai**: Conceptualization, Methodology, Investigation, Writing – original draft, Visualization, Writing – review & editing, Supervision.

