## Supplementary Materials for "Disproportionate Mental Health Risks in Autistic Females: A Rapid Review with Quantitative and Narrative Syntheses"

**Method : search strategy** ____ **2**

**Supplementary Results** ____ **5**

Other MH conditions ____ 5

Transdiagnostic symptoms and pattern of co-occurring conditions ____ 6

Autistic traits in individuals with eating disorders ____ 7

### Method: search strategy

Supplementary Table 1: final search strategy for MEDLINE (October 29, 2024)

| Ovid MEDLINE: Epub Ahead of Print, In-Process & Other Non-Indexed Citations, Ovid MEDLINE® Daily and Ovid MEDLINE® <1946-Present> | | |
|---|---|---|
| 1 | autism spectrum disorder/ | 22754 |
| 2 | autistic disorder/ | 27374 |
| 3 | asperger syndrome/ | 1801 |
| 4 | autis*.ti,kf,hw. or (autis* or ASD).ab. /freq=2 | 73729 |
| 5 | asperger*.ti,kf,hw. or (asperger* or ASD).ab. /freq=2 | 34116 |
| 6 | or/1-5 | 74444 |
| 7 | Women/ or Female/ | 9964305 |
| 8 | (woman or women).ti,kf,hw. or (woman or women).ab. /freq=2 | 782862 |
| 9 | female*.ti,kf,hw. or female*.ab. /freq=2 | 10048325 |
| 10 | (trans* wom?n or trans* female* or transwom* or transfemale* or transfeminine or trans feminine).ti,ab,kf. | 5304 |
| 11 | (trans* m?n or trans* male* or transman* or transmen or transmale* or transmasc* or trans masc*).ti,ab,kf. | 4361 |
| 12 | ((autis* or asperger* or asd) adj3 mother*).ti,ab,kf. | 436 |
| 13 | (sex or sexes or gender*).ti,kf,hw. or (sex or sexes or gender*).ab. /freq=2 | 788531 |
| 14 | or/7-13 | 10294494 |
| 15 | mental disorders/ or exp anxiety disorders/ or exp "disruptive, impulse control, and conduct disorders"/ or exp dissociative disorders/ or exp elimination disorders/ or exp "feeding and eating disorders"/ or exp mood disorders/ or exp motor disorders/ or exp neurocognitive disorders/ or exp neurotic disorders/ or exp paraphilic disorders/ or exp personality disorders/ or exp "schizophrenia spectrum and other psychotic disorders"/ or exp sexual dysfunctions, psychological/ or exp sleep wake disorders/ or exp somatoform disorders/ or exp substance-related disorders/ or exp "trauma and stressor related disorders"/ | 1395161 |
| 16 | exp Psychiatry/ | 112476 |
| 17 | ((comorbid* or co-morbid* or multimorbid* or multi-morbid* or cooccur* or co-occur* or concurrent* or coexist* or coexist* or concomitant or associate* or multiple*) adj3 (mental* or psychiatr*)).ti,kf,hw. | 6680 |

| | | |
|---|---|---|
| 18 | (mood* disorder* or affective* disorder* or anxiety disorder* or depress* or dysthymi* or phobia* or panic* disorder* or obsess* disorder* or compuls* disorder* or OCD or bipolar or bi-polar or personality disorder* or borderline personalit* or manic or mania or psychosis or psychoses or psychotic or schizo* or delusion* or (hear* adj2 voice*)).ti,kf,hw. or (mood* disorder* or affective* disorder* or anxiety disorder* or depress* or dysthymi* or phobia* or panic* disorder* or obsess* disorder* or compuls* disorder* or OCD or bipolar or bi-polar or personality disorder* or borderline personalit* or manic or mania or psychosis or psychoses or psychotic or schizo* or delusion* or (hear* adj2 voice*)).ab. /freq=2 | 825461 |
| 19 | (post-trauma* or posttrauma* or PTSD or complex trauma or developmental trauma or CPTSD).ti,kf,hw. or (post-trauma* or posttrauma* or PTSD or complex trauma or developmental trauma or CPTSD).ab. /freq=2 | 80937 |
| 20 | ((disorder* adj2 eating) or anorex* or bulimi*).ti,kf,hw. or ((disorder* adj2 eating) or anorex* or bulimi*).ab. /freq=2 | 53918 |
| 21 | (suicid* or selfharm* or self-harm* or selfinjur* or self-injur*).ti,kf,hw. or (suicida* or selfharm* or self-harm* or selfinjur* or self-injur*).ab. /freq=2 | 102596 |
| 22 | ((behavio?r* or gambl* or shop* or buy* or spend* or steal* or theft*) adj3 (disorder* or addiction* or compuls* or problem* or pathological*)).ti,kf,hw. or ((behavio?r* or gambl* or shop* or buy* or spend* or steal* or theft*) adj3 (disorder* or addiction* or compuls* or problem* or pathological*)).ab. /freq=2 | 60784 |
| 23 | (Kleptomania* or Trichotillomania or (hair adj3 pull*) or dermatillomania or excoriation or (skin adj3 pick*)).ti,kf,hw. or (Kleptomania* or Trichotillomania or (hair adj3 pull*) or dermatillomania or excoriation or (skin adj3 pick*)).ab. /freq=2 | 2230 |
| 24 | (attention deficit* or ADHD or tourette*).ti,kf,hw. or (attention deficit* or ADHD or tourette*).ab. /freq=2 | 53493 |
| 25 | Emotion* dysregulation.ti,kf,hw. or Emotion* dysregulation.ab. /freq=2 | 2175 |
| 26 | Oppositional defian*.ti,kf,hw. or Oppositional defian*.ab. /freq=2 | 966 |
| 27 | (Conduct adj3 (disorder* or problem* or issue*)).ti,kf,hw. or (Conduct adj3 (disorder* or problem* or issue*)).ab. /freq=2 | 10051 |
| 28 | (Behavio?r* adj3 disorder*).ti,kf,hw. or (Behavio?r* adj3 disorder*).ab. /freq=2 | 37527 |
| 29 | ((externaliz* or externalis* or internalize* or internalis*) adj3 disorder*).ti,ab,kf,hw. | 1721 |
| 30 | (dementia* or alzheimer*).ti,kf,hw. or (dementia* or alzheimer*).ab. /freq=2 | 262430 |

| | | |
|---|---|---|
| 31 | ((substance* or drug* or tobacco or nicotine or alcohol* or cannabis or marijuana or stimulant* or steroid* or analgesic or sedative* or cocaine or hallucinogen* or psilocybin or amphetamine* or methamphetamine* or benzodiazepine* or opioid* or opiate* or heroin or fentanyl or inhalant* or depressant* or barbiturate*) adj3 ("use" or user* or misus* or abus* or disorder* or depend* or addict* or withdraw* or detox* or overdose* or recovery)).ti,kf,hw. or ((substance* or drug* or tobacco or nicotine or alcohol* or cannabis or marijuana or stimulant* or steroid* or analgesic or sedative* or cocaine or hallucinogen* or psilocybin or amphetamine* or methamphetamine* or benzodiazepine* or opioid* or opiate* or heroin or fentanyl or inhalant* or depressant* or barbiturate*) adj3 ("use" or user* or misus* or abus* or disorder* or depend* or addict* or withdraw* or detox* or overdose* or recovery)).ab. /freq=2 | 378287 |
| 32 | SUD.ti,kf,hw. or SUD.ab. /freq=2 | 5482 |
| 33 | ("People who use drugs" or PWUD).ti,ab,kf,hw. | 1777 |
| 34 | ((assist* or supervis* or manag* or treat* or inpatient* or residential*) adj3 (withdraw* or detox*)).ti,ab,kf,hw. | 14010 |
| 35 | ((opioid* or opiate*) adj3 (replacement or substitution or maintenance)).ti,ab,kf,hw. | 7254 |
| 36 | (opioid agonist* adj3 (treatment* or therap*)).ti,ab,kf,hw. | 1591 |
| 37 | ((medica* assisted or opioid assisted) adj3 (treatment* or therap*)).ti,kf,hw. or ((medica* assisted or opioid assisted) adj3 (treatment* or therap*)).ab. /freq=2 | 776 |
| 38 | (naloxone or methodone or buprenorphine or suboxone or sublocade).ti,kf,hw. or (naloxone or methodone or buprenorphine or suboxone or sublocade).ab. /freq=2 | 32231 |
| 39 | (lived experience* or living experience* or PWLE*).ti,ab,kf,hw. | 15489 |
| 40 | or/15-39 [mental health] | 2201263 |
| 41 | 6 and 14 and 40 | 7781 |
| 42 | exp animals/ | 27586104 |
| 43 | exp Animal Experimentation/ | 10608 |
| 44 | exp Models, Animal/ | 664191 |
| 45 | exp Vertebrates/ | 26812379 |
| 46 | or/42-45 | 27588083 |
| 47 | exp Humans/ | 22315435 |
| 48 | exp Human Experimentation/ | 12697 |
| 49 | 46 not (47 or 48) [human filter] | 5272617 |
| 50 | 41 not 49 | 7542 |
| 51 | limit 50 to yr="2004 -Current" | 6294 |

### Supplementary Results

### Other mental health (MH) conditions

A few studies reported sex-stratified findings on other MH conditions, but data remain too limited to draw concrete conclusions.

In a large ClinCom study with children and adolescents (N autism = 1,251), symptoms of *Disruptive Mood Dysregulation Disorder* (DMDD) were present in 43% of the autistic sample, with males exhibiting more symptoms than females (Mayes et al., 2019); in Fombonne et al. (2020), DMDD was diagnosed in 4.6% of autistic adults, with no sex differences. These symptoms overlap with those of *Oppositional Defiant Disorder* (ODD) or *Disruptive Behavior Disorders* broadly, and most participants presenting DMDD also presented ODD, suggesting these conditions may not be phenotypically distinct (Mayes et al., 2019). A PopReg study in autistic children and adolescents (Hsu et al., 2024) identified male sex as a risk factor for *Disruptive Behavior Disorders*. Similarly, a ClinCom study in autistic children also identified male sex as a risk factor for ODD (Salazar et al., 2015). However, this sex-differential risk was not replicated in another ClinCom study in autistic children and adolescents (Albores-Gallo et al., 2017), nor in three PopReg studies in autistic children, adolescents (Dillon et al., 2023), and adults (Fombonne et al., 2020; Jadav & Bal, 2022).

*Conduct disorders* were also more prevalent in autistic male children and adolescents than in females in a PopReg study (Rødgaard et al., 2021). However, no sex differences were reported in PopReg studies with smaller samples of autistic children and adolescents (Dillon et al., 2023; Kerns et al., 2020; Stacy et al., 2014) and adults (Fombonne et al., 2020; Jadav & Bal, 2022; Kentrou et al., 2021), nor in ClinCom studies (Amr et al., 2012; Gobrial, 2019; Salazar et al., 2015).

Sex differences in *Post-Traumatic Stress Disorders (PTSD) or Trauma related disorders* remain underexplored. One PopReg study (Kentrou et al., 2021) indicated that autistic adult females had a 8.2% chance of having a prior diagnosis and 17% of having a co-occurring diagnosis of trauma-related disorder compared to 2.2% and 6.1% in males. One study reported that 52% of autistic male children and adolescents and 67% of autistic females met screening criteria for PTSD, substantially higher than those in non-autistic peers (19% of males and 12% of females); however these sex differences was not statistically significant (Bitsika & Sharpley, 2023). Self-reported symptoms were more severe than parent-reported symptoms (Bitsika & Sharpley, 2023), possibly reflecting limited disclosure or parental awareness of trauma. Another study found higher rates of sexual abuse reported by parents in autistic female children and adolescents (7.1%) than in males (1.4%) (McDonnell et al., 2022).

Geriatric neurological conditions that have a significant impact on MH also lack sufficient sex-stratified research. One PopReg (Croen et al., 2015) and one ClinCom study in people with ID (Tsakanikos et al., 2011) reported higher rates of *dementia* in autistic adult females (up to 4%) compared to males (up to 2%). Rates of dementia were also elevated compared to the non-autistic population (Croen et al., 2015), suggesting possibly increased vulnerability in autistic females specifically. This difference was significant in one study (Tsakanikos et al., 2011) but not investigated in the other (Croen et al., 2015).

### Transdiagnostic symptoms and pattern of co-occurring conditions

Beyond specific diagnoses, several studies have explored transdiagnostic symptoms frequently associated with MH difficulties. For instance, Saez-Suanes et al. (2023) found a relationship between *emotional dysregulation* and anxiety in autistic adults with ID, but significant for females only. They suggested that difficulties in regulating emotions may render autistic females particularly vulnerable to internalizing psychopathology. This is consistent with findings of heightened emotional dysregulation in autistic females compared to males (Weiner et al., 2023; Wieckowski et al., 2020). Furthermore, in autistic adult females, heightened emotional dysregulation was related to higher levels of alexithymia and poorer psychological health, whereas in autistic males, it was associated with heightened autistic traits, poorer physical health, and lower living conditions (Weiner et al., 2023). Emotional dysregulation was also elevated in autistic compared to non-autistic children and adolescents (Wieckowski et al., 2020) and in autistic adult females compared to females with borderline personality disorder (Weiner et al., 2023). In addition, *irritability*, which is part of emotional dysregulation, was heightened in autistic female children and adolescents compared to males in both ClinCom (Graziosi & Perry, 2023) and PopReg studies (Neuhaus et al., 2019; Viscidi et al., 2014). In contrast, *aggression* was rated similarly by teachers in autistic preschool females and males (Gadow et al., 2004), although this may depend on whether it is directed toward others or the self. Indeed, while more autistic adolescent males (26%) than females (11%) were referred to emergency psychiatry for being a danger to others (So et al., 2021), self-injurious behaviors were higher in autistic female children, adolescents and adults (14% in the PopReg study by Martini et al., 2022) than in males (5%) (Cassidy et al., 2018; Maddox et al., 2017; Martini et al., 2022; Steenfeldt-Kristensen et al., 2020). These prevalence rates were higher than those observed in non-autistic individuals, among whom no sex differences were reported (Cassidy et al., 2018; Maddox et al., 2017; Martini et al., 2022).

Some studies investigated broadband *internalizing* and *externalizing disorders* or *symptoms*. A narrative review on adolescents and adults reported heightened rates of internalizing disorders (such as anxiety, depression, ED) in autistic females, while autistic males showed more externalizing problems (Green et al., 2019). A similar trend was observed by professionals who have at least 5 years of experience working with autistic people (Lundin et al., 2021). ClinCom studies also showed heightened internalizing symptomatology in autistic females than males in toddlers (Hartley & Sikora, 2009), children and adolescents (Solomon et al., 2012; Wright et al., 2023). However, other studies found no sex differences in internalizing and/or externalizing symptoms (Holtmann et al., 2007; Jolly et al., 2020; Mayes et al., 2020; Nasca et al., 2020; Neuhaus et al., 2019; Woodman et al., 2016) and one study even found fewer internalizing problems in autistic preschool females compared to males (Prosperi et al., 2020). Comparisons with non-autistic groups also revealed variable findings: while three studies showed higher internalizing (Jamison & Schuttler, 2015; Solomon et al., 2012) and/or externalizing symptoms in autistic females compared to non-autistic females (Jamison & Schuttler, 2015; Woodman et al., 2016), two studies reported fewer internalizing symptoms in autistic than in non-autistic people (Yeung et al., 2024), particularly in females (Woodman et al., 2016). Additionally, a study explored frontal alpha asymmetry (FAA), a neural marker linked to affective style and internalizing-externalizing profiles (Neuhaus et al., 2023). While FAA was associated with internalizing symptoms in the non-autistic sample, it did not significantly correlate with such symptoms in autistic females, raising the possibility that internalizing symptoms in this group may be associated with different neurophysiological mechanisms or may emerge differently over time.

Moreover, the profile of co-occurring condition associations differed by sex: in autistic females co-occurring conditions were more centered around depression, while in males, they were more centered around conduct disorders, impulse control and ADHD (Brown et al., 2020). Some studies, however, found opposite or null results. Autistic male children and adolescents were more likely than females to have one co-occurring diagnosis according to one ClinCom study (Fuca et al., 2023), or multiple according to one PopReg study (Stacy et al., 2014), although the latter also include conditions such as seizure, hearing problem, developmental delay, ADHD, and learning disability. Four ClinCom studies found no sex differences in psychiatric disorders in autistic children (Amr et al., 2012; Memari et al., 2012) and in autistic adolescents with ID (Bradley & Bolton, 2006), or in psychiatric symptoms in autistic children and adolescents (Worley & Matson, 2011).

### Autistic traits in individuals with eating disorders

While ED are highly prevalent in autistic females, the reverse may also be true: two narrative reviews indicated that 4.0-52.5% of females with anorexia nervosa may also be autistic, or exhibit elevated autistic traits (Brown & Stokes, 2020; Westwood & Tchanturia, 2017; see also the studies of Wentz et al., 2005; Zhang et al., 2022). Some authors have proposed a neurodevelopmental etiology for ED, with females being more vulnerable, and anorexia potentially representing a female-specific manifestation of autism (Carpita et al., 2022; Odent, 2010). However, parental reports of early autistic traits in females with ED suggested much lower rates of individuals meeting the autism cut-off (Westwood & Tchanturia, 2017). Several potential explanations have been proposed, such as the possibility that autism characteristics may overlap with ED characteristics, or that parents may under-recognize their child's behaviors as being associated with autism. Whether there is an overlap in characteristics between autism and ED (Nistico et al., 2022), or whether autism is underdiagnosed in females with ED, better screening for autism and clearer distinction of traits are recommended in individuals with anorexia nervosa (Beygui & Cascio, 2022).

**For bibliography, see the main manuscript.**
